# Poor Sleep Health Traits Influence Liking of Sweet Foods and Sugary Food Intake: A UK Biobank Study

**DOI:** 10.64898/2026.02.15.26346360

**Authors:** Phoebe SC Hui, Calista D Touw, Surabhi Bhutani, Liang-Dar Hwang

## Abstract

Poor sleep is linked to consumption of sugary foods/beverages and high neural responsivity to palatable food cues. Yet, whether hedonic liking for sweet taste explains these associations remains unclear. We examined cross-sectional associations of five sleep traits (chronotype, sleep duration, insomnia frequency, snoring, daytime dozing) and a composite sleep score with sweet food liking, and total and free sugar intake in 76,734 UK Biobank participants (39-72 years, 56.3% female). Models adjusted for age, sex, ethnicity, socioeconomic deprivation, and body mass index (Bonferroni-corrected α=0.0025). Evening chronotype, more frequent insomnia and daytime dozing, and lower composite sleep score were associated with higher sweet food liking. Associations with intake were stronger for free than total sugar. Evening chronotype was associated with higher free sugar intake (g/day: β=1.523, 1.309-1.737; g/1000 kcal: β=0.450, 0.361-0.538), and daytime dozing showed a dose-response (dozing often vs never/rarely: g/day β=6.307, 4.631-7.983). Snoring was associated with higher absolute (but not energy-adjusted) free sugar intake. A healthier sleep score was associated with lower free sugar intake (g/day β=-2.193 [−2.464 to −1.922]; g/1000 kcal β=-0.691 [−0.804 to −0.579]) but higher energy-adjusted total sugar intake (β=0.633 [0.485-0.781]). Mediation analyses indicated sweet liking accounted for 15%-91% of several sleep trait and free sugar intake associations (indirect effects p<0.001). Poorer sleep health, particularly evening chronotype and daytime sleepiness, was associated with greater sweet liking and higher free sugar intake, with sweet liking partially mediating associations between sleep traits and sugar consumption. Sweet-taste liking may represent an underexamined pathway linking sleep/circadian disruption to free sugar intake.

## INTRODUCTION

Our sense of taste plays a critical role in food selection and long-term dietary patterns.^1^ Humans show an innate attraction to sweet-tasting foods from early life,^1^ and higher sweet liking has been associated with greater consumption of energy-dense foods and sugar-sweetened beverages, contributing to obesity and related metabolic risks.^2,3^ Emerging, though inconsistent, evidence also suggests that obesity may be linked to altered sweet taste perception and stronger hedonic responses to sweet stimuli,^4,5^ supporting potentially bidirectional relationships between sweet-taste processing, diet, and adiposity.

Sleep and circadian characteristics are also closely linked to diet and body weight regulation. Experimental sleep restriction studies show that curtailed sleep increases intake of high-energy foods, sweet snacks, and sugar-sweetened beverages, and can shift eating toward greater carbohydrate intake and snacking outside regular mealtimes.^6–8^ Observational studies associate short or fragmented sleep, insomnia symptoms, habitual snoring, and daytime sleepiness/dozing with poorer overall diet quality, including higher intakes of added sugars and saturated fat and lower fibre and fruit/vegetable intake.^8–10^ Chronotype, preferred timing of the sleep-wake cycle, may further shape diet and weight via circadian differences in meal timing and food choice.^11–13^ Evening chronotypes, characterized by later and more irregular sleep and meal timing, have been linked to higher free sugar-rich food and beverage intake, and lower fruit and vegetable consumption.^14–16^ Proposed mechanisms include changes in appetite-regulating hormones, heightened reward sensitivity to palatable foods, and impaired executive control, which together may promote hedonic eating.^17–19^ Neurobehavioral studies further support stronger reward-related neural responses to energy-dense food cues following sleep disruption or circadian misalignment.^19–22^

Despite these converging findings, the role of taste hedonics in sleep-diet relationships remains poorly understood. Sleep and diet share many determinants and may influence one another bi-directionally, so observed associations can reflect causal effects, shared vulnerabilities, or both. Sweet-taste liking is associated with obesity and sugar-rich dietary patterns, yet few studies have examined whether multiple sleep traits (e.g. chronotype, insomnia, snoring, daytime dozing) relate to sweet liking and free sugar intake within the same analytic framework.^23^ Experimental evidence suggests that even partial sleep loss can alter perceived sweetness and increase hedonic responses to sweet stimuli,^23^ raising the possibility that sleep disruption increases the reward valuation of sweetness and, in turn, free sugar consumption. Free sugars (added sugars and sugars in sweetened beverages) are a priority target in dietary guidelines, and are more consistently associated with adverse cardiometabolic outcomes.^24,25^ However, population-scale evidence integrating multidimensional sleep measures, sweet liking, and free sugar intake remains limited. It is unclear whether sweet taste liking statistically accounts for observed sleep–sugar associations. Although recent work suggests that sleep loss may preferentially influence hedonics, most studies are small laboratory experiments that do not link changes in liking/preference to habitual dietary intake.^23^ Similarly, while sleep extension interventions have reported reductions in free sugar intake,^26^ they typically do not assess concurrent changes in sweet-taste hedonics, leaving the behavioral pathway linking sleep and sugar consumption unresolved.

The present study uses the UK Biobank cohort to address these gaps in a large, well-characterized population sample. We aimed to 1) test associations of chronotype (extreme morning vs extreme evening), sleep duration, insomnia frequency, snoring, daytime sleepiness, and a composite sleep score with liking of sweet foods, 2) test associations between these sleep measures and intake of total and free sugars; and 3) evaluate whether sweet food liking mediates the associations between sleep-related traits and total and free sugars intake. To our knowledge, this is among the first large population studies to examine multiple sleep traits, hedonic liking of sweet taste, and free vs total sugar intake in a single analytic framework. By integrating multidimensional sleep measures with hedonic sweet-food liking and distinguishing free from total sugars within a single analytic framework, this study aims to clarify which sleep dimensions show the most coherent associations with sugar intake and whether sweet liking is a plausible taste-related correlate of sleep-diet relationships, informing priorities for future longitudinal and experimental tests of taste-related pathways linking sleep/circadian disruption with sugar consumption and obesity risk.

## METHODS

### Data and Study Population

UK Biobank (UKB) is a population-based cohort of >500,000 adults aged 40-69 years, recruited across the United Kingdom between 2006 and 2010. Comprehensive biomedical data, including lifestyle characteristics and health metrics, were collected from all participants at baseline and at follow-up assessments.^27^

Among 501,936 participants (application 53641) with data available on the UKB Research Analysis Platform at the time of data extraction and analysis (27 October 2025). Total 76,734 participants with complete baseline data for all five sleep traits, covariates, and at least one valid outcome measure (sweet liking and/or sugar intake) were included in the present study. Participants were included in each analysis if they had valid data for the specific outcome variable (liking or intake measure), resulting in varying sample sizes across models (reported in **Table 1**). The flow of participants through each stage of inclusion and exclusion is shown in **Figure 1**.

**Figure 1.**
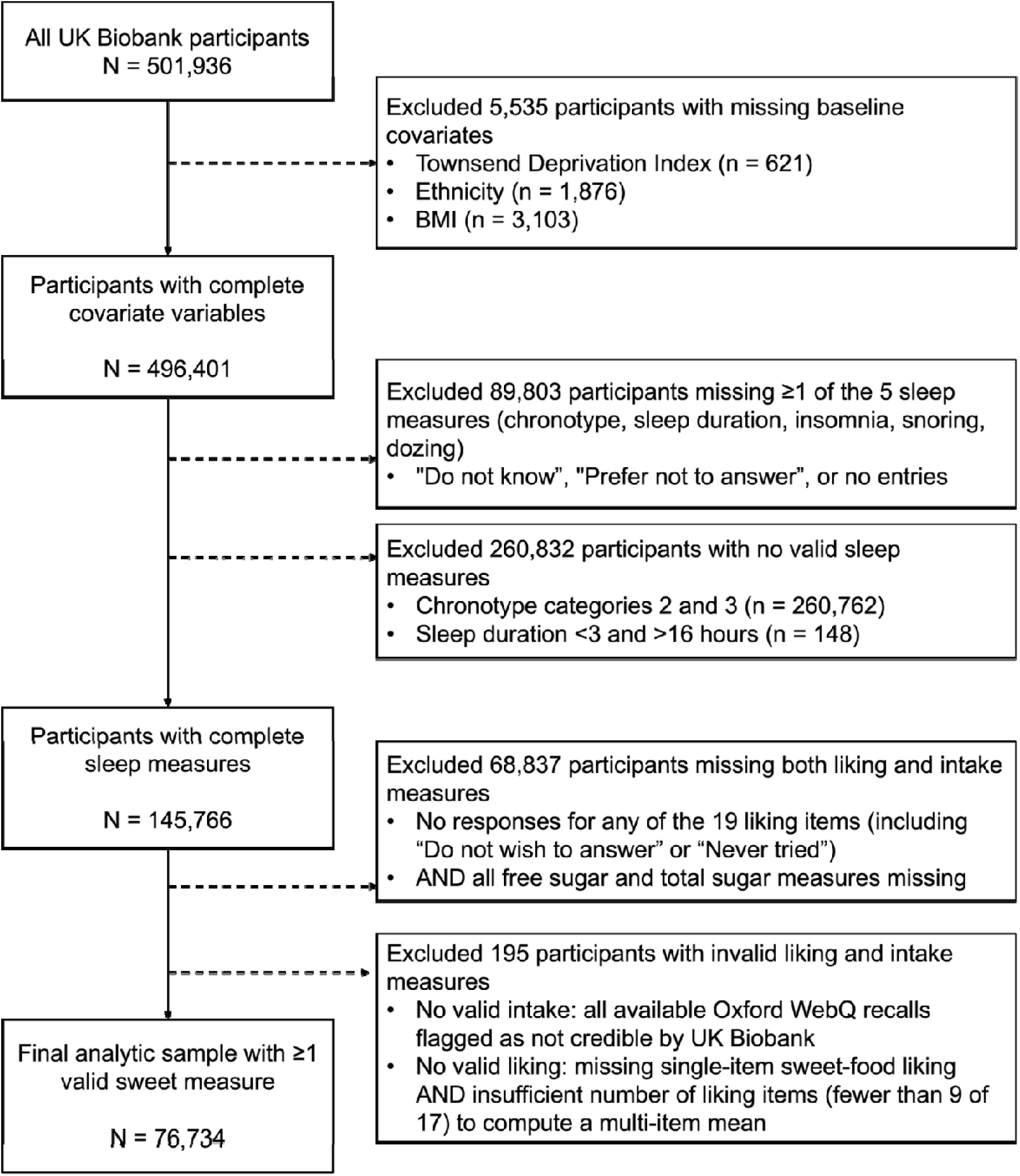
Summary of participants included and excluded from the analysis. Flowchart illustrating the sequential inclusion and exclusion of participants. Participants were excluded only if they had missing baseline covariates, missing sleep data, ambiguous chronotype responses, implausible sleep duration values, or absence of any credible sweet liking or sugar intake data. Participants with valid data for at least one outcome were retained and contributed to outcome-specific analyses.

**Table 1.** Sample characteristics.

| Characteristic | Total sample population (N = 76,734) |  |
| --- | --- | --- |
| Age (years, mean ± SD) | 56.1 ± 7.8 | |
| Sex (N [%]) |  |  |
| Female | 43,212 (56.3) |  |
| Male | 33,522 (43.7) |  |
| TDI (mean ± SD) | -1.6 ± 2.9 | |
| BMI (kg/m<sup>2</sup>, mean ± SD) | 27.2 ± 4.8 | |
| Ethnic background (N [%]) |  |  |
| White | 73,238 (95.4) |  |
| Asian or Asian British | 1,202 (1.6) |  |
| Black or Black British | 971 (1.3) |  |
| Mixed | 505 (0.7) |  |
| Chinese | 217 (0.3) |  |
| Other ethnic group | 601 (0.8) |  |
| Chronotype (N [%]) |  |  |
| Definitely morning | 56,455 (73.6) |  |
| Definitely evening | 20,279 (26.4) |  |
| Sleep duration (hours/day, mean ± SD) | 7.1 ± 1.1 | |
| Insomnia (N [%]) |  |  |
| Never/rarely | 20,606 (26.9) |  |
| Sometimes | 34,636 (45.1) |  |
| Usually | 21,492 (28.0) |  |
| Snoring (N [%]) |  |  |
| No | 48,794 (63.6) |  |
| Yes | 27,940 (36.4) |  |
| Daytime dozing (N [%]) |  |  |
| Never/rarely | 59,167 (77.1%) |  |
| Sometimes | 15,390 (20.1%) |  |
| Often | 2,177 (2.8%) |  |
| Composite sleep score (mean ± SD) | 3.1 ± 1.1 | |
| Single-item sweet food liking (mean ± SD) | 5.9 ± 2.2 | (n = 51,620) |
| Average sweet liking (multi-item) (mean ± SD) | 5.2 ± 1.1 | (n = 51,659) |
| Total sugar intake |  |  |
| in grams (mean ± SD) | 124.5 ± 49.7 | (n = 60,429) |
| in grams per 1000 kcal (mean ± SD) | 62.0 ± 19.4 | (n = 60,423) |
| Free sugar intake |  |  |
| in grams (mean ± SD) | 59.7 ± 36.2 | (n = 60,429) |
| in grams per 1000 kcal (mean ± SD) | 28.8 ± 14.7 | (n = 60,423) |

*Abbreviations:* SD, standard deviation; TDI, Townsend Deprivation Index; BMI, body mass index.

UKB received approval from the Northwest Multi-Centre Research Ethics Committee (MREC) as a Research Tissue Bank (RTB) in 2011 (11/NW/0382), 2016 (16/NW/0274) and 2021 (21/NW/0157). All participants provided consent prior to participating in the UKB.

### Assessment of Sleep Variables

Five baseline sleep traits were derived from the UKB touchscreen questionnaire (2006-2010): chronotype, sleep duration, insomnia frequency, snoring, and daytime dozing. Chronotype was assessed using the following question: “Do you consider yourself to be *1*) definitely a ‘morning’ person, *2*) more a ‘morning’ than an ‘evening’ person, *3*) more an ‘evening’ than a ‘morning’ person, or *4*) definitely an ‘evening’ person.” In line with previous analyses, ^28,29^ we only included individuals in categories 1 and 4 for robust phenotyping of chronotype.

Sleep duration was obtained with the question “About how many hours sleep do you get in every 24 hours?” Sleep duration was reported as hours per 24 h and values <3 or >16 h/day were set to missing and excluded from analyses. Insomnia was assessed by asking the participants about the frequency of trouble with falling asleep at night and awakenings.

Responses included: “never/rarely”; “sometimes”, or “usually”. Snoring was assessed by self-report of complaints about snoring (yes/no). Daytime sleepiness/dozing was assessed as the likelihood of falling asleep during the daytime, with responses ranging from “never/rarely” to “often”. For all five sleep variables, responses of “Do not know” or “Prefer not to answer” were set to missing.

To summarize overall sleep health, we calculated a composite sleep score as a pragmatic summary of five self-reported sleep behaviors using previously published low-risk criteria.^30,31^ Each sleep behavior was coded as low-risk (1) or high-risk (0), summing the scores (range 0-5; higher scores indicate healthier sleep). Low risk behaviors were defined as: sleep duration of 7-9 hours per day, ‘definitely morning’ chronotype, never or rarely experiencing insomnia, no self-reported snoring, and never or rarely experiencing daytime dozing. All other responses were classified as high-risk. This score is not a validated clinical instrument; therefore, we also report associations for each individual sleep trait (including mutually adjusted models) to ensure findings are not driven by the composite definition. The score was analyzed as a continuous predictor and interpreted as an overall indicator of sleep health rather than a validated scale.

### Assessment of Sweet Food Liking

Sweet food liking was derived from the UKB Food Preference Questionnaire administered in 2019 to participants who had agreed to being recontacted. Participants rated their liking for each of the 140 food items and other behaviors, such as physical activity, on a 9-point hedonic scale (1=extremely dislike, 9=extremely like) Responses “Never tried” and “Do not wish to answer” were coded as missing.

We used two complementary measures of sweet food liking: 1) the single-item “sweet foods” liking rating; and 2) the mean liking score across 17 pre-specified individual sweet food items (items listed in **Supplementary Table 1)**, excluding the single-item ‘sweet foods’ liking rating. For the multi-item liking score, participants were required to have a valid response (1-9) for ≥9 of 17 items (>50%) to ensure reliable individual-level liking estimates. This threshold was implemented to minimize the risk of averages being disproportionately influenced by a small subset of items among individuals with substantial missing data, especially given that some items showed skewed distributions (e.g., ∼50% reported extreme dislikes for fizzy drinks). Notably, 99.9% of respondents met this threshold.

### Assessment of Diet Intake Variables

Diet data were collected using the Oxford WebQ (www.ceu.ox.ac.uk/research/oxford-webq) web-based 24-hour Dietary Recall, completed up to five times between April 2009 and June 2012. The Oxford WebQ has been validated against an interviewer-administered 24-hour dietary recall, producing a mean Spearman correlation coefficient for macronutrients of 0.62 (range 0.54-0.69). ^32^ Free sugars intake (g/day) and total sugars intake (g/day) were extracted from the UK Nutrient Databank linked within UKB. These values represent absolute sugar intake. Oxford WebQ entries flagged by UKB as implausible energy intake were excluded (>4,302 kcal for females; >4,790 kcal for males). To account for differences in total energy intake, we also extracted total energy intake (kcal/day) and estimated free and total sugar intake as grams of sugar per 1000 kcal (g/1000kcal), hereafter referred to as energy-adjusted sugar intake. Mean intakes were calculated across all valid WebQ assessments if a participant had multiple valid entries.

### Assessment of Covariates

Covariates including age at recruitment, self-reported sex (male/female), the Townsend Deprivation Index (TDI) an area-based proxy measure for socioeconomic status (SES) provided in the UKB directly, self-reported ethnic background, and body mass index (BMI) were documented through touchscreen questionnaires, verbal interview records, and physical measures at baseline. ^33^ Ethnicity was re-coded to top-level ethnic groups (e.g., British to White, Indian to Asian, Caribbean to Black) for analysis.

### Statistical analysis

Baseline characteristics of the 76,734 participants were summarized as mean (standard deviation [SD]) or number (percentage). Linear regression models estimated the association of each sleep measure with i) liking for sweet foods (single-item), ii) average sweet liking (multi-item mean), and iii) total and free sugar intake, expressed as absolute (grams per day) and energy-adjusted (grams per 1000kcal) values.

Given robust associations between sweet liking and total and free sugar intake (**Supplementary Table 2**), we used structural equational modelling (SEM) to examine whether sweet liking mediates associations between sleep behaviors and sugar intake (**Figure 2**). Within each SEM model, we estimated: (1) the indirect effect (sleep ➔ liking ➔ intake, calculated as the product of paths a and b); (2) the direct effect (sleep ➔ intake after accounting for the mediator, path c’); (3) the total effect (sum of indirect and direct effects), and (4) and the proportion explained by the indirect effect [(indirect/total)×100].

**Figure 2.**
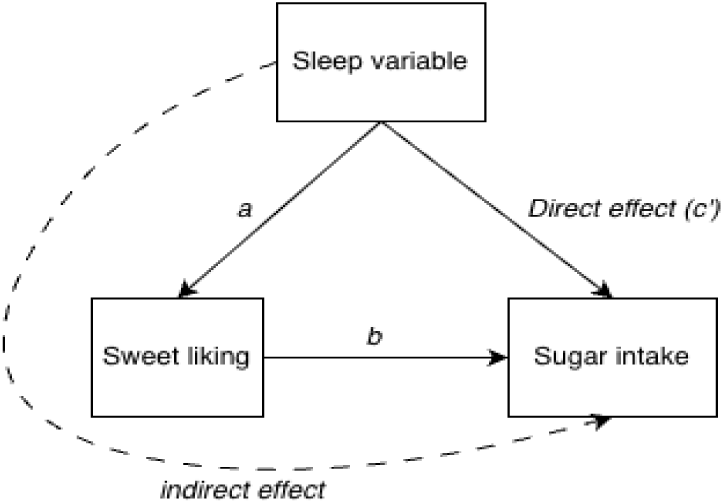
Mediation process pathway for a model of sleep variables and sugar intake. The straight arrows represent the relationship between variables and suggest a causal relation from the base to the head of the arrow. Direct effect is the effect of sleep variables on sugar intake (total sugar and free sugar in g and g/1000kcal) while controlling for sweet liking (single-item sweet food liking rating and average sweet liking rating for sweet foods). Indirect effect is the effect of sleep variables on sugar intake through sweet liking. The significance of total effects (defined as the sum of indirect and direct effects) and the individual relationships (represented by the estimates) between the three variables were used to determine whether sugar intake was mediated by liking for sweet foods.

Due to varying completeness across liking and intake measures, SEM sample sizes ranged from 35,328 to 35,359 participants, depending on the liking measure (single-item “sweet foods” or multi-item average) and intake metric (g/day or g/1000 kcal). SEM was conducted using the lavaan R package ^34^.

All models, including linear regression and SEM, were adjusted for all covariates. Sleep duration and composite sleep score were treated as continuous variables, while chronotype and snoring were analyzed as binary variables. Insomnia and daytime dozing were recorded as categorical variables with three levels and modelled as continuous variables (0/1/2) in SEM, as dummy coding for categorical variables would produce separate estimates for each level, complicating the interpretation of indirect effects. For individual sleep traits, we fitted both univariate models (each sleep behavior examined separately, adjusted only for standard covariates) and mutually adjusted models (all five sleep behaviors included simultaneously as predictors). Only univariate models were conducted for the composite sleep score, as the score itself represents a proxy for the combined contribution of all five sleep behaviors.

Correlation matrices were constructed for the sleep predictors and for the sweet liking with sugar intake outcomes to estimate the effective number of independent tests using matSpD ^35^ (https://research.qut.edu.au/sgel/software/matspd-local-version/). The effective number of independent tests was determined to be 5 independent sleep predictors × 4 independent sweet liking or intake outcomes, resulting in Bonferroni correction for 20 independent tests and an alpha significance threshold of 0.05/20 = 0.0025. Analyses were conducted in R (v4.4.0) on the UK Biobank Research Analysis Platform.

## RESULTS

**Table 1** summarizes the characteristics of the full sample (N = 76,734) of individuals with complete data on food liking or food intake and all sleep and covariates. The mean age of our sample was 56.1 ± 7.8 years, predominantly female (56.3%), and identified as white (95.4%). The mean BMI was 27.2 ± 4.8 kg/m^²^. With respect to sleep traits, 26.4% reported being an Evening chronotype, one-third reported “usually” having insomnia (28.0%), 36.4% reported “yes” to complaints about snoring (36.4%), and one-fifth reported experiencing daytime dozing sometimes (20.1%) or often (2.8%). Participants reported an average sleep duration of 7.1 ± 1.1 hours/night. Mean composite sleep score was 3.1 ± 1.1 (range 0 to 5, higher scores indicate a healthier sleep pattern).

### Association of sleep traits with sweet food liking

Associations between sleep traits and sweet food liking are presented in **Table 2** and summarized in **Supplementary Figure 1**. In multivariable-adjusted models (Bonferroni-corrected α ≤ 0.0025), evening chronotype was associated with higher single-item liking rating for sweet foods (β=0.067, 95% CI 0.053-0.082) and higher average liking of pre-specified sweet items (β=0.044, 0.037-0.052), compared with morning chronotype. Daytime dozing showed the strongest dose-response pattern, such that compared with never/rarely dozing, dozing sometimes and often were associated with higher single-item sweet liking (β=0.276, 0.228-0.325 and β=0.468, 0.349-0.586) and higher average sweet liking (β=0.169, 0.145-0.194 and β=0.228, 0.168-0.288). Insomnia frequency was also positively associated with both liking measures. In contrast, sleep duration was not associated with sweet liking after multiple-testing correction, and snoring was associated with higher average sweet liking (β=0.063, 0.042-0.083, p=4.83×10^-^C) but not the single-item rating (p=4.58×10^-^³). Most associations were robust to mutually adjusting for all five sleep behaviors, except for the association between usually experiencing insomnia and average sweet liking, which attenuated after adjustment (**Supplementary Table 3)**.

**Table 2.**
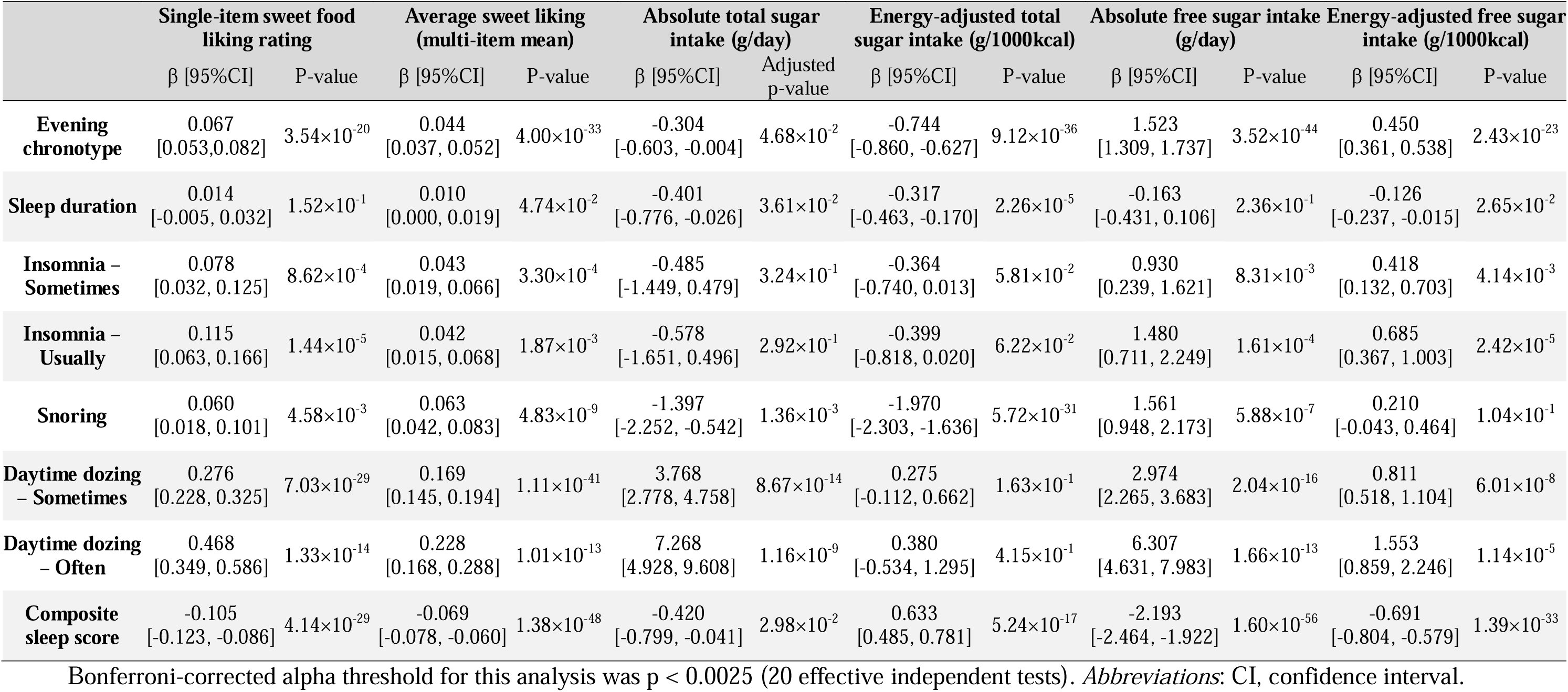
Association between sleep traits with liking for sweet foods and sugar intake.

|  | Single-item sweet food liking rating |  | Average sweet liking (multi-item mean) |  | Absolute total sugar intake (g/day) |  | Energy-adjusted total sugar intake (g/1000kcal) |  | Absolute free sugar intake (g/day) |  | Energy-adjusted free sugar intake (g/1000kcal) |  |
| --- | --- | --- | --- | --- | --- | --- | --- | --- | --- | --- | --- | --- |
| | $\beta$ [95% CI] | P-value | $\beta$ [95% CI] | P-value | $\beta$ [95% CI] | Adjusted p-value | $\beta$ [95% CI] | P-value | $\beta$ [95% CI] | P-value | $\beta$ [95% CI] | P-value |
| **Evening chronotype** | 0.067<br>[0.053, 0.082] | $3.54 \times 10^{-20}$ | 0.044<br>[0.037, 0.052] | $4.00 \times 10^{-33}$ | -0.304<br>[-0.603, -0.004] | $4.68 \times 10^{-2}$ | -0.744<br>[-0.860, -0.627] | $9.12 \times 10^{-36}$ | 1.523<br>[1.309, 1.737] | $3.52 \times 10^{-44}$ | 0.450<br>[0.361, 0.538] | $2.43 \times 10^{-23}$ |
| **Sleep duration** | 0.014<br>[-0.005, 0.032] | $1.52 \times 10^{-1}$ | 0.010<br>[0.000, 0.019] | $4.74 \times 10^{-2}$ | -0.401<br>[-0.776, -0.026] | $3.61 \times 10^{-2}$ | -0.317<br>[-0.463, -0.170] | $2.26 \times 10^{-5}$ | -0.163<br>[-0.431, 0.106] | $2.36 \times 10^{-1}$ | -0.126<br>[-0.237, -0.015] | $2.65 \times 10^{-2}$ |
| **Insomnia – Sometimes** | 0.078<br>[0.032, 0.125] | $8.62 \times 10^{-4}$ | 0.043<br>[0.019, 0.066] | $3.30 \times 10^{-4}$ | -0.485<br>[-1.449, 0.479] | $3.24 \times 10^{-1}$ | -0.364<br>[-0.740, 0.013] | $5.81 \times 10^{-2}$ | 0.930<br>[0.239, 1.621] | $8.31 \times 10^{-3}$ | 0.418<br>[0.132, 0.703] | $4.14 \times 10^{-3}$ |
| **Insomnia – Usually** | 0.115<br>[0.063, 0.166] | $1.44 \times 10^{-5}$ | 0.042<br>[0.015, 0.068] | $1.87 \times 10^{-3}$ | -0.578<br>[-1.651, 0.496] | $2.92 \times 10^{-1}$ | -0.399<br>[-0.818, 0.020] | $6.22 \times 10^{-2}$ | 1.480<br>[0.711, 2.249] | $1.61 \times 10^{-4}$ | 0.685<br>[0.367, 1.003] | $2.42 \times 10^{-5}$ |
| **Snoring** | 0.060<br>[0.018, 0.101] | $4.58 \times 10^{-3}$ | 0.063<br>[0.042, 0.083] | $4.83 \times 10^{-9}$ | -1.397<br>[-2.252, -0.542] | $1.36 \times 10^{-3}$ | -1.970<br>[-2.303, -1.636] | $5.72 \times 10^{-31}$ | 1.561<br>[0.948, 2.173] | $5.88 \times 10^{-7}$ | 0.210<br>[-0.043, 0.464] | $1.04 \times 10^{-1}$ |
| **Daytime dozing – Sometimes** | 0.276<br>[0.228, 0.325] | $7.03 \times 10^{-29}$ | 0.169<br>[0.145, 0.194] | $1.11 \times 10^{-41}$ | 3.768<br>[2.778, 4.758] | $8.67 \times 10^{-14}$ | 0.275<br>[-0.112, 0.662] | $1.63 \times 10^{-1}$ | 2.974<br>[2.265, 3.683] | $2.04 \times 10^{-16}$ | 0.811<br>[0.518, 1.104] | $6.01 \times 10^{-8}$ |
| **Daytime dozing – Often** | 0.468<br>[0.349, 0.586] | $1.33 \times 10^{-14}$ | 0.228<br>[0.168, 0.288] | $1.01 \times 10^{-13}$ | 7.268<br>[4.928, 9.608] | $1.16 \times 10^{-9}$ | 0.380<br>[-0.534, 1.295] | $4.15 \times 10^{-1}$ | 6.307<br>[4.631, 7.983] | $1.66 \times 10^{-13}$ | 1.553<br>[0.859, 2.246] | $1.14 \times 10^{-5}$ |
| **Composite sleep score** | -0.105<br>[-0.123, -0.086] | $4.14 \times 10^{-29}$ | -0.069<br>[-0.078, -0.060] | $1.38 \times 10^{-48}$ | -0.420<br>[-0.799, -0.041] | $2.98 \times 10^{-2}$ | 0.633<br>[0.485, 0.781] | $5.24 \times 10^{-17}$ | -2.193<br>[-2.464, -1.922] | $1.60 \times 10^{-56}$ | -0.691<br>[-0.804, -0.579] | $1.39 \times 10^{-33}$ |

Bonferroni-corrected alpha threshold for this analysis was $p < 0.0025$ (20 effective independent tests). *Abbreviations:* CI, confidence interval.

A healthier composite sleep score was associated with lower sweet liking (single-item: β=−0.105, −0.123 to −0.086; average: β=-0.069, −0.078 to −0.060). For both individual sleep behaviors and the composite sleep score, results for average sweet liking remained consistent in sensitivity analyses restricted to participants with complete responses for all 17 items (**Supplementary Table 3**), suggesting that the ≥9-item threshold provided a robust measure of average sweet liking.

### Association of sleep traits with free and total sugar intake

Associations between sleep traits and dietary sugar intake are presented in **Table 2** and summarized in **Supplementary Figure 1**. Overall, associations were more consistent for free sugars than for total sugars and differed for absolute (g/day) versus energy-adjusted (g/1000 kcal) measures.

For free sugars, evening chronotype was associated with higher free sugar intake in both absolute (g/day: β=1.523, 1.309-1.737) and energy-adjusted models (g/1000 kcal: β=0.450, 0.361-0.538). Daytime dozing showed a dose-response pattern, with higher free sugar intake among those dozing often versus never/rarely (g/day β=6.307, 4.631-7.983; g/1000 kcal β=1.553, 0.859-2.246). Snoring was associated with higher absolute free sugar intake (β=1.561, 0.948-2.173; p=5.88×10^-7^) but was not associated with energy-adjusted free sugar intake. Insomnia (“usually”) was associated with higher free sugar in both absolute and energy-adjusted models. No association was observed for insomnia (“sometimes”) or sleep duration.

Associations with total sugar were weaker and depended on energy adjustment.

Snoring was associated with lower total sugar intake in both absolute (g/day β=-1.397, −2.252 to −0.542; p=1.36×10^-^³) and energy-adjusted total sugar intake models (β=-1.970, −2.303 to - 1.636; p=5.72×10C³¹). Evening chronotype and longer sleep duration were associated with lower total sugar intake in the energy-adjusted models but not in the absolute models.

Daytime dozing was associated with higher total sugar intake in the absolute models but not in the energy-adjusted models. No association was observed for insomnia. Results were consistent in models mutually adjusted for all five sleep traits.

In analyses of composite sleep score, a higher score (healthier sleep) was associated with lower free sugar intake in both absolute (g/day β=-2.193, −2.464 to −1.922) and energy-adjusted models (g/1000 kcal β=-0.691, −0.804 to −0.579). It was associated with higher energy-adjusted total sugar intake (β=0.633, 0.485–0.781; p = 5.24×10^−17^) but not with absolute total sugar intake.

### Mediation analysis

Effect estimates from mediation analysis are presented in **Table 3**. For free sugar outcomes, both sweet liking measures showed significant indirect effects for several sleep traits and the composite sleep score, indicating that sweet liking statistically accounted for part of the observed sleep–free sugar associations. Evening chronotype, higher insomnia frequency, snoring, and more frequent daytime dozing were all associated with higher free sugar intake (both absolute [g/day] and energy-adjusted [g/1000kcal] intake), with 15-91% of these effects operating through increased sweet liking (p < 0.001 for all indirect effects). The composite sleep score showed similar patterns, with sweet liking accounting for 15-30% of the inverse associations between healthier sleep score and free sugar intake (**Supplementary Figure 2**). These patterns were largely consistent across both liking measures (single ‘sweet food’ item or average sweet liking) and remained significant after adjusting for other sleep behaviors (**Supplementary Table 4**).

**Table 3.**
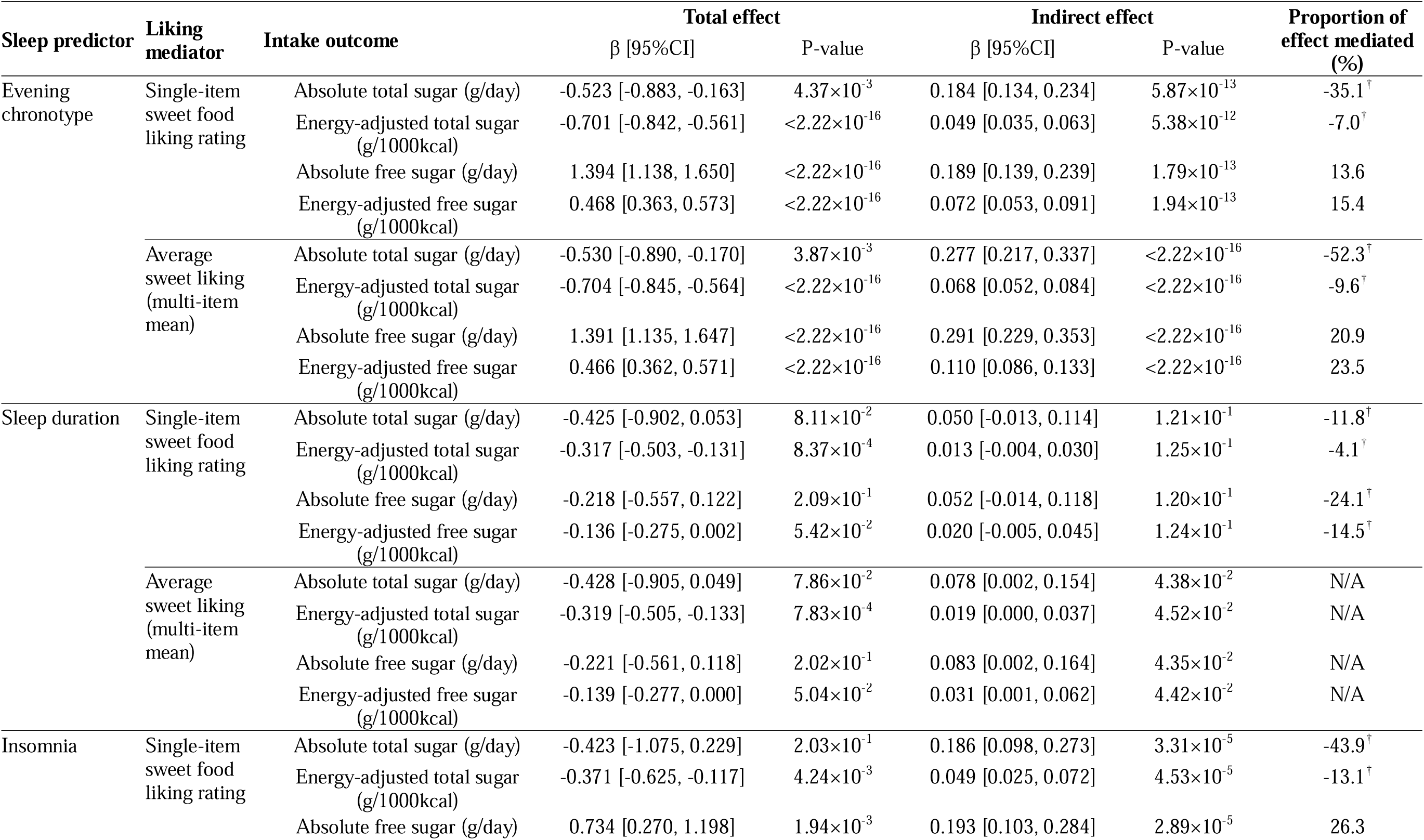
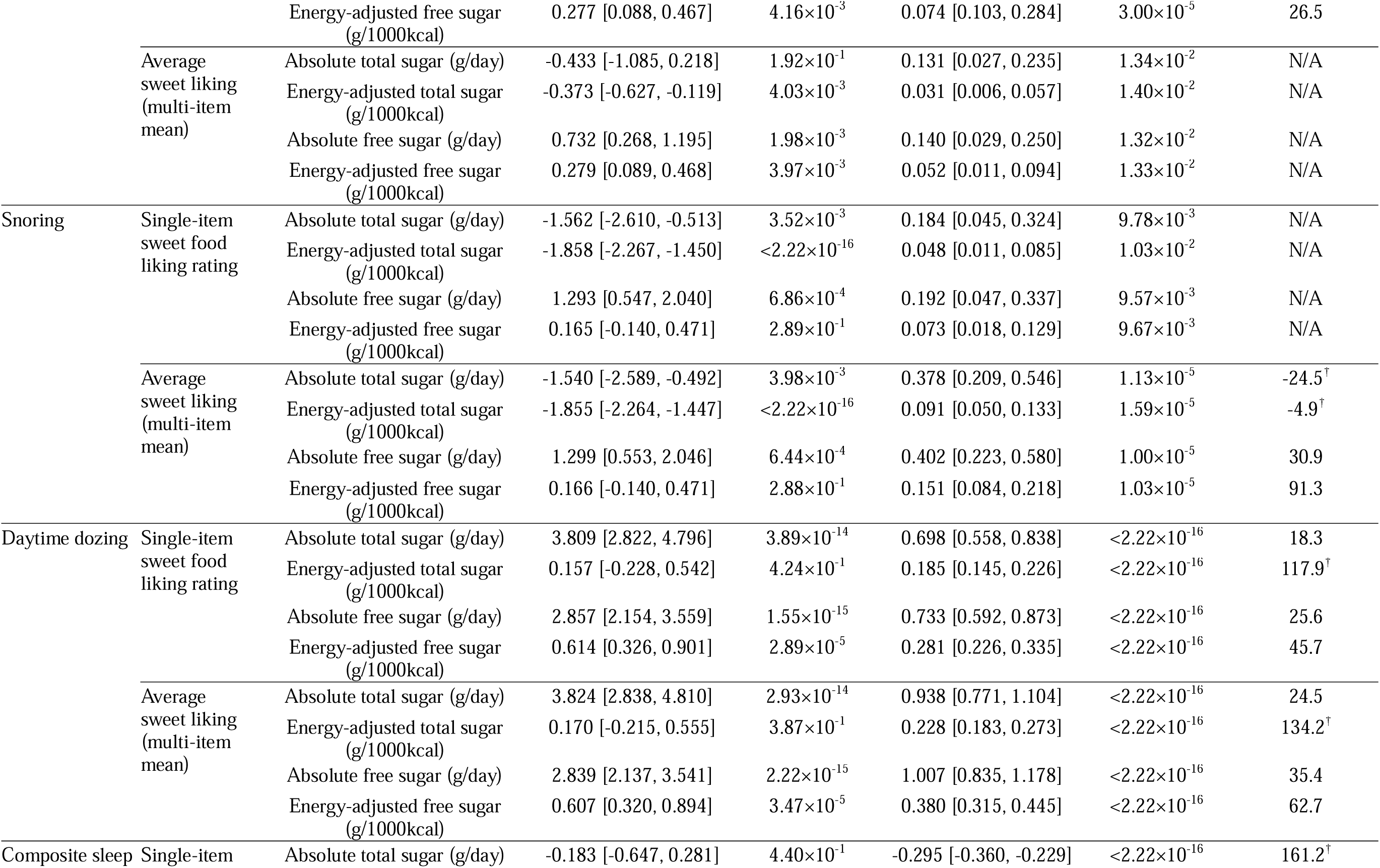
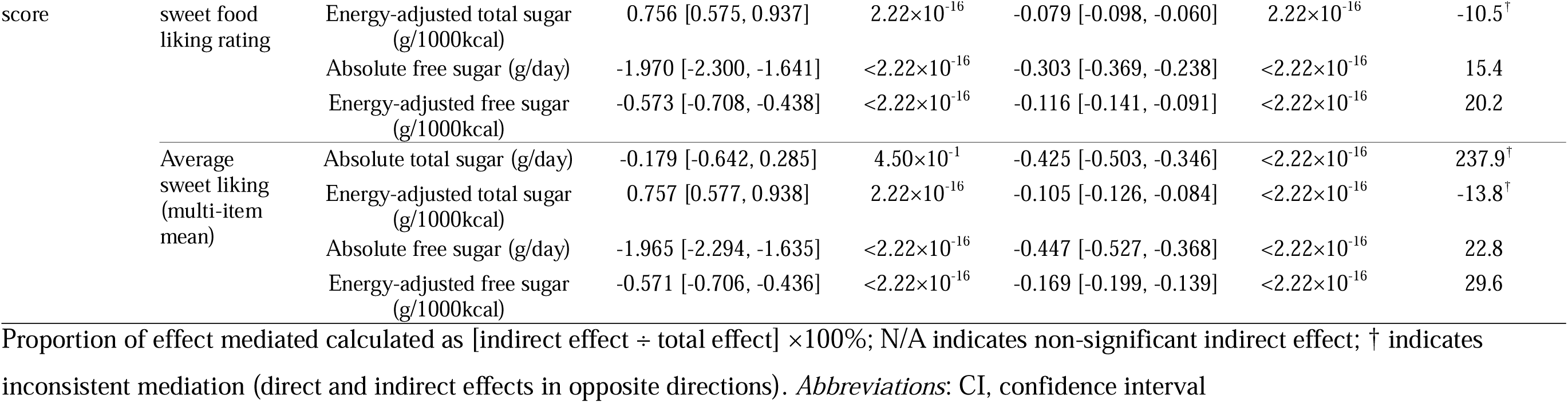
Association between sleep traits and sugar intake with sweet food liking as a mediator.

For total sugar outcomes, mediation patterns were more complex. Evening chronotypes, insomnia, and snoring showed inconsistent mediation, where the indirect effect through sweet liking operated in the opposite direction to the direct effect (sleep trait to intake). Daytime dozing also showed inconsistent mediation for energy-adjusted total sugar intake but demonstrated significant mediation (18.4-24.5% mediated) for absolute total sugar intake. The composite sleep score similarly showed inconsistent mediation for total sugar intake. Sleep duration showed either inconsistent mediation or no significant indirect effects through sweet liking for any outcome, despite showing a negative association with energy-adjusted total sugar intake in linear regression models.

## DISCUSSION

Using the large UKB cohort, we examined associations of multiple self-reported sleep traits with hedonic liking for sweet foods and dietary intake of total and free sugars. Several dimensions of poorer sleep health, including evening chronotype, more frequent insomnia, snoring, daytime dozing, and a lower composite sleep score, were associated with higher sweet food liking. These traits also showed more consistent associations with free sugar intake than with total sugars, with particularly robust patterns for evening chronotype and daytime dozing across both absolute and energy-adjusted outcomes. In contrast, associations with total sugar intake were weaker and sometimes inverse when expressed relative to energy intake. Mediation analyses suggested that sweet liking statistically accounted for part of several observed sleep and free sugar intake associations, whereas patterns for total sugar were inconsistent. Sleep duration showed little evidence of association with sweet liking or free sugar intake, suggesting that qualitative sleep dimensions may be more relevant than sleep quantity alone.

Our findings align with experimental and epidemiological evidence linking sleep with taste-related hedonics. A recent systematic review concluded that shorter or poor sleep quality is most consistently associated with higher preferred sweetness concentration, whereas evidence for sweet taste sensitivity/detection thresholds is mixed and often null.^23^ Experimental sleep-curtailment studies similarly report higher preferred sucrose (or sweetener) concentrations and higher liking ratings following acute sleep loss, with limited evidence of changes in detection threshold.^36^ Population data in Chinese adults further suggests that poor self-reported sleep quality (including insomnia, daytime sleepiness, and short sleep) is associated with altered taste perception.^37^ Extending this literature beyond sleep duration/acute deprivation, we observed robust associations for multiple qualitative sleep dimensions and for an aggregate sleep health score in mid- to late adulthood, independent of key sociodemographic factors and BMI. Given that taste hedonics reflect multiple influences (e.g., genetics, learned exposure, sociocultural factors),^38,39^ even modest shifts in sweet liking may be relevant at the population level for free sugar intake, which is concentrated in ultra-processed foods and sugar-sweetened beverages.

Evening chronotype and daytime dozing showed the most consistent associations with both sweet liking and free sugar intake. This pattern is consistent with observational evidence that evening chronotypes are reported to have less healthy dietary profiles than morning types, including higher consumption of energy-dense, sugar-rich foods and lower fruit and vegetable intake.^14–16^ Social jetlag and circadian misalignment, correlates of evening chronotype, have also been linked to higher intakes of sugar-sweetened beverages and added sugars among individuals with later or more irregular sleep-wake timing. In young adults, late chronotypes exhibit a greater desire (wanting) for highly palatable high-fat foods compared with earlier chronotypes.^40^ Although wanting and liking are distinct constructs, they often co-occur and interact,^40^ consistent with our observation of higher sweet liking among evening types. While daytime dozing and excessive daytime sleepiness are less studied in relation to diet, our observed associations are biologically and behaviorally plausible. Daytime sleepiness may increase reliance on rewarding, sugar-rich snacks and beverages to combat fatigue, aligning with experimental evidence that sleep loss increases the selection of highly palatable, carbohydrate-rich foods.^41,42^ The parallel associations with both sweet liking and free sugar intake suggest that hedonic valuation of sweet foods may be one correlate of sleepiness-related differences in dietary behavior. Snoring, a proxy for sleep-disordered breathing, was also associated with higher multi-item sweet liking and higher absolute free sugar intake despite lower total sugar intake. Prior population studies indicate that poor sleep quality, often operationalized using insomnia symptoms, snoring, daytime sleepiness, and sleep duration, is associated with altered taste perception.^37^ In obstructive sleep apnoea, disturbances in taste and smell have been reported and may relate to intermittent hypoxia and inflammation,^43^ providing a biological context for taste-related differences among individuals with sleep-disordered breathing symptoms.

Across sleep traits, associations with dietary intake were more coherent for free sugars than for total sugars. This aligns with evidence that free sugars, particularly from sugar-sweetened beverages and discretionary foods, are more consistently linked to adverse cardiometabolic outcomes than intrinsic sugars from whole foods.^24^ Our composite sleep score, capturing multidimensional sleep health, further suggested that healthier sleep is associated with lower sweet liking and lower free sugar intake but higher total sugar relative to calories. This pattern may reflect differences in sugar sources, such as relatively higher intake of intrinsic sugars from fruits and dairy, alongside lower intake of free sugar-rich foods and beverages.^44,45^ Consistent with this interpretation, indirect-effect patterns were often inconsistent for total sugar outcomes, likely because total sugars combine intrinsic and free sugar sources.

Our mediation analyses suggested that sweet liking statistically accounted for part of several sleep–free sugar associations. These findings are consistent with mechanistic evidence that sleep restriction can heighten activity in reward-related brain regions and reduce prefrontal control during response to food cues, increasing the salience of sweet, energy-dense foods.^19–22^ They are also consistent with evidence that sleep loss alters appetite-regulating hormones such as ghrelin and leptin, which may promote intake of sweets and snacks.^17,46^ For total sugar intake, mediation patterns were often inconsistent, with indirect effects via sweet liking opposing direct associations between sleep traits and sugar energy adjusted sugar intake. This likely reflects the fact that total sugars combine both free and intrinsic sugars. For example, a sleep trait that increases intake of free-sugar-rich foods but reduces consumption of fruits and dairy could lower total sugar per 1000 kcal despite higher free sugar intake. Overall, our findings highlight the importance of separating free from intrinsic sugars when interpreting indirect-effect models.

Strengths of this study include the large sample size, evaluation of multiple sleep traits and a composite sleep score, and use of both single- and multi-item measures of sweet liking. Distinguishing free from total sugars and considering both absolute and energy-adjusted intakes allowed a nuanced characterization of sleep–diet associations. The mediation analysis further quantified the proportion of sleep–diet associations that may operate through sweet liking. Limitations should also be acknowledged. The observational design precludes causal inference and does not exclude bidirectional relationships between sleep, taste, and diet.^8^ Sleep traits and taste liking were self-reported and may be subject to misclassification and reporting bias. Although we adjusted for key sociodemographic and anthropometric covariates, residual confounding by factors such as mental health, shift work, and medication use is possible. ^37^ Additionally, the UKB cohort has a low response rate (∼5%) and is predominantly White, which may limit generalizability to more diverse populations.

Despite these limitations, our findings have important implications for interventions targeting sleep health and dietary quality. Approaches addressing evening chronotype, insomnia symptoms, daytime sleepiness, and sleep-breathing may be relevant for reducing free sugar intake, particularly if paired with strategies that reduce exposure to highly sweet foods and beverages and potentially reshape hedonic liking. Chrono-nutrition approaches that align meal timing and composition with circadian biology may be especially relevant for evening chronotypes.^47,48^ Future longitudinal and experimental studies incorporating objective sleep measures, repeated dietary assessments, and direct measures of taste function and food reward are needed to clarify directionality and test whether improving sleep influences sweet liking and free sugar intake.^23^

## CONCLUSION

To our knowledge, this is one of the first large cohort studies to quantify associations between multiple sleep traits, sweet food liking, and free sugar intake. Several sleep-related traits were associated with higher sweet liking and higher free sugar intake, with indirect-effect models suggesting that sweet-taste liking is an underexamined correlate of sleep–diet relationships and underscoring the importance of distinguishing free from total sugars. Future longitudinal and experimental studies are needed to test whether targeting sleep health and sweet-taste liking can reduce free sugar intake and related metabolic risk.

## Data Availability

All data produced in the present study are available upon reasonable request to the authors. Individual‑level UK Biobank data are available via application to UK Biobank (https://www.ukbiobank.ac.uk/).

## Acknowledgements

This research has been conducted using the UK Biobank Resource under Application Number 53641. We would like to thank the UK Biobank and all its participants. Open access publishing facilitated by The University of Queensland as part of the Wiley-University of Queensland agreement via the Council of Australian University Liberians.

## Data availability statement

Data can be accessed through UK Biobank (https://www.ukbiobank.ac.uk/).

