## Supplementary Materials for "Poor Sleep Health Traits Influence Liking of Sweet Foods and Sugary Food Intake: A UK Biobank Study"

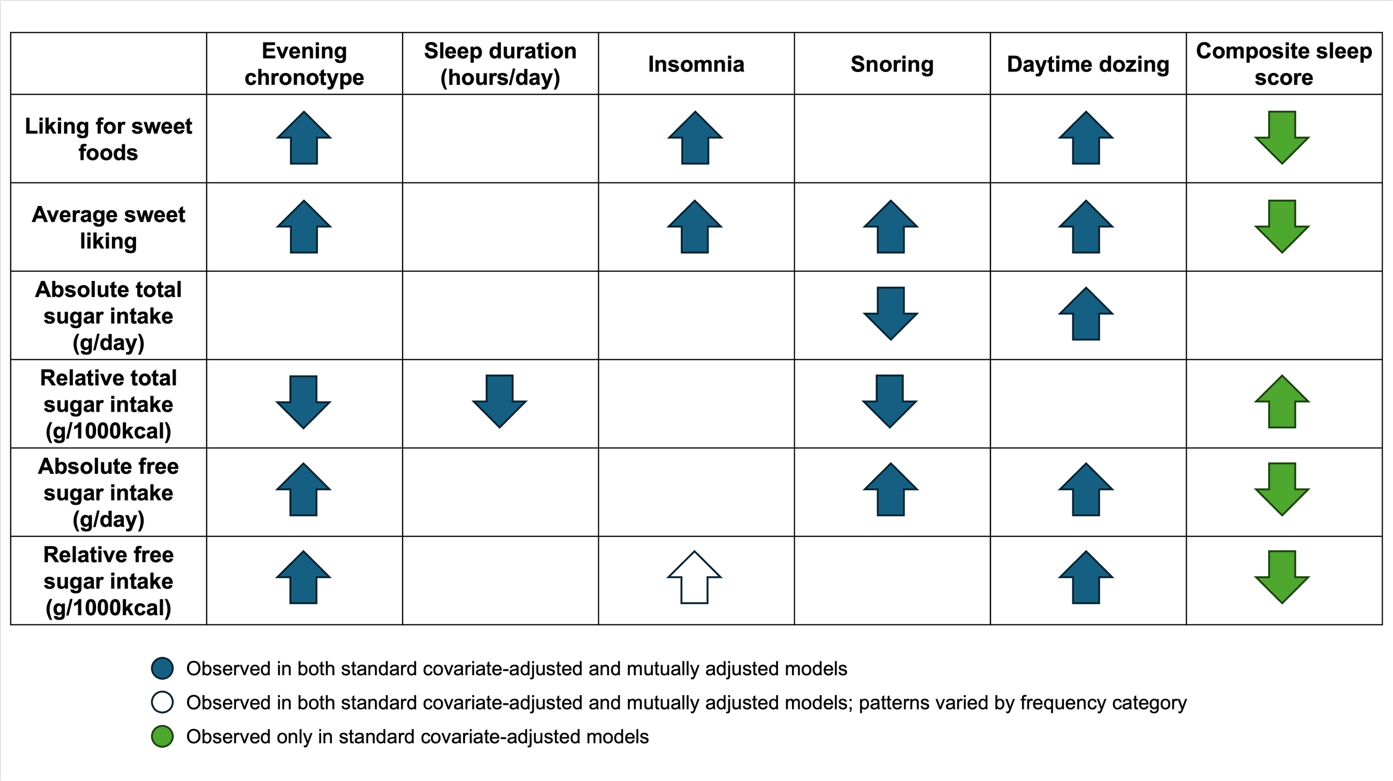

**Supplementary Figure 1. Summary of associations between sleep variables with sweet liking and sugar intake measures.** Arrows indicate the direction of associations between sleep predictors and outcome: upward arrows indicate positive associations; downward arrows indicate negative associations. Only associations with p <0.0025 (Bonferroni-corrected threshold) are shown. All models are adjusted for age, sex, SES, BMI, and ethnicity. Color coding indicates whether associations were observed in standard covariate-adjusted models only (green), or in both standard and mutually adjusted models with all five sleep behaviors (blue). White arrows indicate associations significant in both model specifications but with patterns that vary by frequency category (Sometimes vs Usually for insomnia; see Table 2 for category-specific estimates). For the composite sleep score, mutually adjusted models were not performed as the score itself integrates all five sleep behaviors, with higher scores reflecting healthier overall sleep patterns.

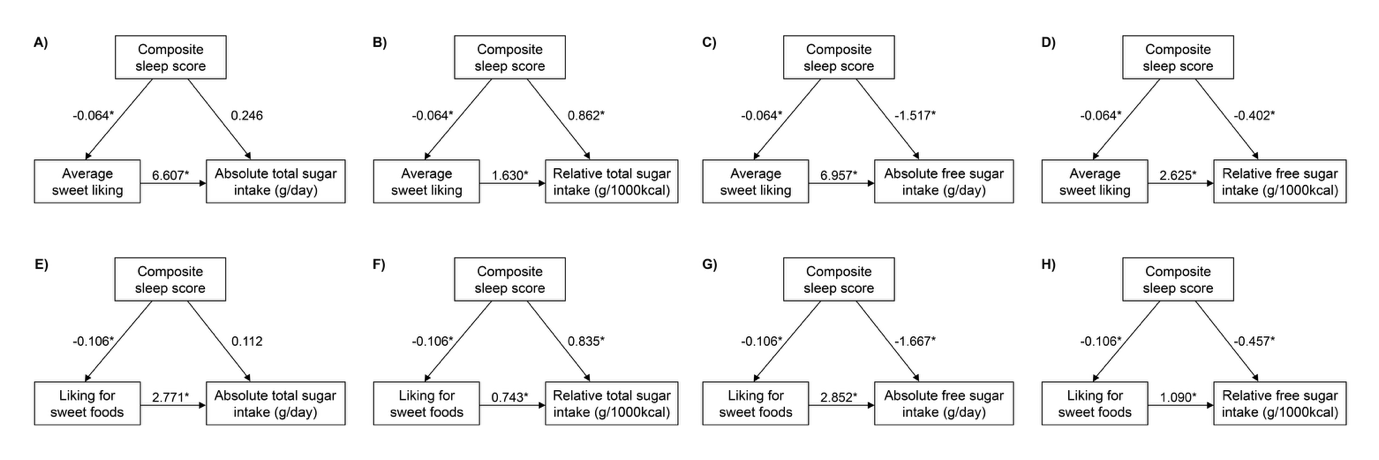

**Supplementary Figure 2. Associations between composite sleep score and sugar intake mediated by sweet liking.** Path coefficients (unstandardized) from structural equation models showing single-item ‘liking for sweet foods’ (A-D) and average sweet liking (E-H) as mediators for absolute total sugar intake (A & E), relative total sugar intake (B & F), absolute free sugar intake (C & G), and relative free sugar intake (D & H). All models adjusted for age, sex, SES, BMI, and ethnicity. * p < 0.0025 (Bonferroni-adjusted threshold).

**Supplementary Table 1.** Items included in 'average sweet liking' calculations

| **Item** | **Trait ID** |
| --- | --- |
| Apple juice | p20602 |
| Biscuits | p20615 |
| Cake | p20631 |
| Cake icing | p20632 |
| Cereal/granola bar | p20635 |
| Cheesecake | p20636 |
| Coffee with sugar | p20643 |
| Dark chocolate | p20652 |
| Diet fizzy drinks | p20653 |
| Ice cream | p20679 |
| Jam | p20680 |
| Marizpan | p20688 |
| Milk chocolate | p20691 |
| Regular fizzy drinks | p20710 |
| Sweet coffee house drinks | p20731 |
| Tea with sugar | p20734 |

**Supplementary Table 2.** Associations of sweet liking with total and free sugar intake (Oxford WebQ) in UK Biobank.

| **Outcome (Y)** | **Covariates** | **Sample size** | **β** | **SE** | **P-value** | **R^2^ (%)** | **Adj R^2^ (%)** | **Cov Adj R^2^ (%)** | **ΔAdj R² attributable to liking (%)** |
| --- | --- | --- | --- | --- | --- | --- | --- | --- | --- |
| **Average sweet liking (multi-item mean)** | | | | | | | | | |
| Free sugar (g/1000kcal) | Standard covariates | 35359 | 2.648 | 0.063 | 0.00x10^300^* | 6.57 | 6.54 | 2.18 | 4.36 |
|  | + 5 sleep traits | 35359 | 2.621 | 0.063 | 0.00x10^300^* | 6.73 | 6.69 | 2.43 | 4.26 |
|  | + composite sleep score | 35359 | 2.625 | 0.063 | 0.00x10^300^* | 6.66 | 6.63 | 2.41 | 4.22 |
| Free sugar (g/day) | Standard covariates | 35362 | 7.042 | 0.153 | 0.00x10^300^* | 11.25 | 11.22 | 5.52 | 5.70 |
|  | + 5 sleep traits | 35362 | 6.919 | 0.153 | 0.00x10^300^* | 11.54 | 11.50 | 6.03 | 5.47 |
|  | + composite sleep score | 35362 | 6.957 | 0.153 | 0.00x10^300^* | 11.46 | 11.43 | 5.91 | 5.52 |
| Total sugar (g/1000kcal) | Standard covariates | 35359 | 1.581 | 0.086 | 1.49x10^-75^* | 3.64 | 3.62 | 2.41 | 1.20 |
|  | + 5 sleep traits | 35359 | 1.655 | 0.086 | 1.73x10^-82^* | 4.29 | 4.24 | 2.91 | 1.33 |
|  | + composite sleep score | 35359 | 1.630 | 0.086 | 5.10x10^-80^* | 3.88 | 3.85 | 2.52 | 1.33 |
| Total sugar (g/day) | Standard covariates | 35362 | 6.593 | 0.218 | 4.07x10^-198^* | 4.69 | 4.66 | 1.99 | 2.68 |
|  | + 5 sleep traits | 35362 | 6.592 | 0.219 | 4.51x10^-197^* | 4.91 | 4.87 | 2.16 | 2.71 |
|  | + composite sleep score | 35362 | 6.607 | 0.219 | 3.15x10^-198^* | 4.69 | 4.66 | 1.99 | 2.67 |
| **Single item sweet food liking** | | | | | | | | | |
| Free sugar (g/1000kcal) | Standard covariates | 35328 | 1.101 | 0.032 | 9.54x10^-258^* | 5.07 | 5.04 | 2.18 | 2.87 |
|  | + 5 sleep traits | 35328 | 1.087 | 0.032 | 1.86x10^-250^* | 5.27 | 5.22 | 2.43 | 2.80 |
|  | + composite sleep score | 35328 | 1.090 | 0.032 | 1.47x10^-252^* | 5.19 | 5.16 | 2.41 | 2.75 |
| Free sugar (g/day) | Standard covariates | 35331 | 2.891 | 0.078 | 1.36x10^-297^* | 9.46 | 9.43 | 5.52 | 3.91 |
|  | + 5 sleep traits | 35331 | 2.832 | 0.078 | 2.43x10^-285^* | 9.82 | 9.77 | 6.03 | 3.75 |
|  | + composite sleep score | 35331 | 2.852 | 0.078 | 6.82x10^-290^* | 9.72 | 9.69 | 5.91 | 3.78 |
| Total sugar (g/1000kcal) | Standard covariates | 35328 | 0.724 | 0.043 | 1.06x10^-62^* | 3.48 | 3.45 | 2.41 | 1.04 |
|  | + 5 sleep traits | 35328 | 0.752 | 0.043 | 1.46x10^-67^* | 4.10 | 4.05 | 2.91 | 1.14 |
|  | + composite sleep score | 35328 | 0.743 | 0.043 | 5.09x10^-66^* | 3.70 | 3.67 | 2.52 | 1.15 |
| Total sugar (g/day) | Standard covariates | 35331 | 2.768 | 0.110 | 5.11x10^-138^* | 3.94 | 3.91 | 1.99 | 1.92 |
|  | + 5 sleep traits | 35331 | 2.759 | 0.110 | 1.08x10^-136^* | 4.15 | 4.11 | 2.16 | 1.95 |
|  | + composite sleep score | 35331 | 2.771 | 0.110 | 6.21x10^-138^* | 3.94 | 3.91 | 1.99 | 1.91 |

Standard covariates include sex, age, body mass index, socioeconomic status (Townsend Deprivation Index), and ethnicity.
*p < 0.0083 (Bonferroni-corrected threshold for 6 independent tests [2 effective independent predictors × 3 effective independent outcome as per matSpD])
Columns: Adj_r^2^, adjusted R^2^ of the full regression model; Cov_adj_r^2^, adjusted R^2^ of the covariate-only model; ΔAdj R² attributable to liking () is variance attributed to liking predictor

**Supplementary Table 3.** Association of sweet food liking and sugar intake outcomes on sleep predictors.

| **Outcome (Y)** | **Models** | **Sample size** | **β** | **SE** | **P-value** | **R^2^ (%)** | **Adj R^2^ (%)** | **Cov Adj R^2^ (%)** | | **ΔAdj R²**  **(%)** |
| --- | --- | --- | --- | --- | --- | --- | --- | --- | --- | --- |
| **Chronotype (Evening vs Morning)** | | | | | | | | | | |
| Average sweet liking (multi-item mean) | Standard covariates | 51659 | 0.044 | 0.004 | 4.00x10-^33^* | 3.23 | 3.21 | 2.94 | | 0.27 |
|  | + 5 sleep traits | 51659 | 0.044 | 0.004 | 1.84x10^-32^* | 3.72 | 3.69 | 2.94 | | 0.75 |
| Single item sweet food liking | Standard covariates | 51620 | 0.067 | 0.007 | 3.54x10^-20^* | 2.17 | 2.15 | 1.99 | | 0.16 |
|  | + 5 sleep traits | 51620 | 0.066 | 0.007 | 1.09x10^-19^* | 2.54 | 2.51 | 1.99 | | 0.51 |
| Total sugar (g/day) | Standard covariates | 60429 | -0.304 | 0.153 | 4.68x10^-02^ | 2.01 | 1.99 | 1.99 | | 0.00 |
|  | + 5 sleep traits | 60429 | -0.308 | 0.153 | 4.39x10^-02^ | 2.18 | 2.16 | 1.99 | | 0.17 |
| Total sugar (g/1000kcal) | Standard covariates | 60423 | -0.744 | 0.060 | 9.12x10^-36^* | 2.68 | 2.66 | 2.41 | | 0.25 |
|  | + 5 sleep traits | 60423 | -0.737 | 0.059 | 3.39x10^-35^* | 2.94 | 2.91 | 2.41 | | 0.50 |
| Free sugar (g/day) | Standard covariates | 60429 | 1.523 | 0.109 | 3.52x10^-44^* | 5.84 | 5.82 | 5.52 | | 0.30 |
|  | + 5 sleep traits | 60429 | 1.513 | 0.109 | 1.14x10^-43^* | 6.05 | 6.03 | 5.52 | | 0.51 |
| Free sugar (g/1000kcal) | Standard covariates | 60423 | 0.450 | 0.045 | 2.43x10^-23^* | 2.35 | 2.34 | 2.18 | | 0.16 |
|  | + 5 sleep traits | 60423 | 0.449 | 0.045 | 3.07x10^-23^* | 2.45 | 2.43 | 2.18 | | 0.25 |
| **Sleep duration (per hour)** | | | | | | | | | | |
| Average sweet liking (multi-item mean) | Standard covariates | 51659 | 0.010 | 0.005 | 4.74x10^-02^ | 2.97 | 2.95 | 2.94 | | 0.01 |
|  | + 5 sleep traits | 51659 | 0.013 | 0.005 | 6.62x10^-03^ | 3.72 | 3.69 | 2.94 | | 0.75 |
| Single item sweet food liking | Standard covariates | 51620 | 0.014 | 0.009 | 1.52x10^-01^ | 2.02 | 2.00 | 1.99 | | 0.00 |
|  | + 5 sleep traits | 51620 | 0.025 | 0.010 | 9.85x10^-03^ | 2.54 | 2.51 | 1.99 | | 0.51 |
| Total sugar (g/day) | Standard covariates | 60429 | -0.401 | 0.191 | 3.61x10^-02^ | 2.01 | 1.99 | 1.99 | | 0.01 |
|  | + 5 sleep traits | 60429 | -0.425 | 0.198 | 3.23x10^-02^ | 2.18 | 2.16 | 1.99 | | 0.17 |
| Total sugar (g/1000kcal) | Standard covariates | 60423 | -0.317 | 0.075 | 2.26x10^-05^* | 2.45 | 2.44 | 2.41 | | 0.03 |
|  | + 5 sleep traits | 60423 | -0.330 | 0.077 | 1.96x10^-05^* | 2.94 | 2.91 | 2.41 | | 0.50 |
| Free sugar (g/day) | Standard covariates | 60429 | -0.163 | 0.137 | 2.36x10^-01^ | 5.54 | 5.52 | 5.52 | | 0.00 |
|  | + 5 sleep traits | 60429 | -0.076 | 0.142 | 5.90x10^-01^ | 6.05 | 6.03 | 5.52 | | 0.51 |
| Free sugar (g/1000kcal) | Standard covariates | 60423 | -0.126 | 0.057 | 2.65x10^-02^ | 2.20 | 2.18 | 2.18 | | 0.01 |
|  | + 5 sleep traits | 60423 | -0.082 | 0.059 | 1.61x10^-01^ | 2.45 | 2.43 | 2.18 | | 0.25 |
| **Insomnia (Sometimes vs Never/rarely)** | | | | | | | | | | |
| Average sweet liking (multi-item mean) | Standard covariates | 51659 | 0.043 | 0.012 | 3.30x10^-04^* | 2.99 | 2.96 | 2.94 | | 0.02 |
|  | + 5 sleep traits | 51659 | 0.043 | 0.012 | 3.06x10^-04^* | 3.72 | 3.69 | 2.94 | | 0.75 |
| Single item sweet food liking | Standard covariates | 51620 | 0.078 | 0.024 | 8.62x10^-04^* | 2.05 | 2.03 | 1.99 | | 0.03 |
|  | + 5 sleep traits | 51620 | 0.079 | 0.024 | 8.20x10^-04^* | 2.54 | 2.51 | 1.99 | | 0.51 |
| Total sugar (g/day) | Standard covariates | 60429 | -0.485 | 0.492 | 3.24x10^-01^ | 2.00 | 1.99 | 1.99 | | 0.00 |
|  | + 5 sleep traits | 60429 | -0.707 | 0.493 | 1.51x10^-01^ | 2.18 | 2.16 | 1.99 | | 0.17 |
| Total sugar (g/1000kcal) | Standard covariates | 60423 | -0.364 | 0.192 | 5.81x10^-02^ | 2.43 | 2.42 | 2.41 | | 0.00 |
|  | + 5 sleep traits | 60423 | -0.492 | 0.192 | 1.03x10^-02^ | 2.94 | 2.91 | 2.41 | | 0.50 |
| Free sugar (g/day) | Standard covariates | 60429 | 0.930 | 0.352 | 8.31x10^-03^ | 5.56 | 5.54 | 5.52 | | 0.02 |
|  | + 5 sleep traits | 60429 | 0.937 | 0.352 | 7.83x10^-03^ | 6.05 | 6.03 | 5.52 | | 0.51 |
| Free sugar (g/1000kcal) | Standard covariates | 60423 | 0.418 | 0.146 | 4.14x10^-03^ | 2.22 | 2.20 | 2.18 | | 0.03 |
|  | + 5 sleep traits | 60423 | 0.410 | 0.146 | 4.91x10^-03^ | 2.45 | 2.43 | 2.18 | | 0.25 |
| **Insomnia (Usually vs Never/rarely)** | | | | | | | | | | |
| Average sweet liking (multi-item mean) | Standard covariates | 51659 | 0.042 | 0.013 | 1.87x10^-03^* | 2.99 | 2.96 | 2.94 | | 0.02 |
|  | + 5 sleep traits | 51659 | 0.039 | 0.014 | 5.16x10^-03^ | 3.72 | 3.69 | 2.94 | | 0.75 |
| Single item sweet food liking | Standard covariates | 51620 | 0.115 | 0.026 | 1.44x10^-05^* | 2.05 | 2.03 | 1.99 | | 0.03 |
|  | + 5 sleep traits | 51620 | 0.109 | 0.027 | 6.63x10^-05^* | 2.54 | 2.51 | 1.99 | | 0.51 |
| Total sugar (g/day) | Standard covariates | 60429 | -0.578 | 0.548 | 2.92x10^-01^ | 2.00 | 1.99 | 1.99 | | 0.00 |
|  | + 5 sleep traits | 60429 | -1.321 | 0.566 | 1.97x10^-02^ | 2.18 | 2.16 | 1.99 | | 0.17 |
| Total sugar (g/1000kcal) | Standard covariates | 60423 | -0.399 | 0.214 | 6.22x10^-02^ | 2.43 | 2.42 | 2.41 | | 0.00 |
|  | + 5 sleep traits | 60423 | -0.725 | 0.221 | 1.02x10^-03^* | 2.94 | 2.91 | 2.41 | | 0.50 |
| Free sugar (g/day) | Standard covariates | 60429 | 1.480 | 0.392 | 1.61x10^-04^* | 5.56 | 5.54 | 5.52 | | 0.02 |
|  | + 5 sleep traits | 60429 | 1.163 | 0.405 | 4.07x10^-03^ | 6.05 | 6.03 | 5.52 | | 0.51 |
| Free sugar (g/1000kcal) | Standard covariates | 60423 | 0.685 | 0.162 | 2.42x10^-05^* | 2.22 | 2.20 | 2.18 | | 0.03 |
|  | + 5 sleep traits | 60423 | 0.555 | 0.168 | 9.36x10^-04^* | 2.45 | 2.43 | 2.18 | | 0.25 |
| **Snoring (Yes vs No)** | | | | | | | | | | |
| Average sweet liking (multi-item mean) | Standard covariates | 51659 | 0.063 | 0.011 | 4.83x10^-09^* | 3.02 | 3.00 | 2.94 | | 0.06 |
|  | + 5 sleep traits | 51659 | 0.051 | 0.011 | 1.56x10^-06^* | 3.72 | 3.69 | 2.94 | | 0.75 |
| Single item sweet food liking | Standard covariates | 51620 | 0.060 | 0.021 | 4.58x10^-03^ | 2.03 | 2.01 | 1.99 | | 0.01 |
|  | + 5 sleep traits | 51620 | 0.041 | 0.021 | 5.34x10^-02^ | 2.54 | 2.51 | 1.99 | | 0.51 |
| Total sugar (g/day) | Standard covariates | 60429 | -1.397 | 0.436 | 1.36x10^-03^* | 2.02 | 2.00 | 1.99 | | 0.02 |
|  | + 5 sleep traits | 60429 | -1.670 | 0.437 | 1.33x10^-04^* | 2.18 | 2.16 | 1.99 | | 0.17 |
| Total sugar (g/1000kcal) | Standard covariates | 60423 | -1.970 | 0.170 | 5.72x10^-31^* | 2.64 | 2.63 | 2.41 | | 0.21 |
|  | + 5 sleep traits | 60423 | -1.976 | 0.170 | 4.45x10^-31^* | 2.94 | 2.91 | 2.41 | | 0.50 |
| Free sugar (g/day) | Standard covariates | 60429 | 1.561 | 0.312 | 5.88x10^-07^* | 5.57 | 5.56 | 5.52 | | 0.04 |
|  | + 5 sleep traits | 60429 | 1.345 | 0.313 | 1.70x10^-05^* | 6.05 | 6.03 | 5.52 | | 0.51 |
| Free sugar (g/1000kcal) | Standard covariates | 60423 | 0.210 | 0.129 | 1.04x10^-01^ | 2.20 | 2.18 | 2.18 | | 0.00 |
|  | + 5 sleep traits | 60423 | 0.162 | 0.129 | 2.12x10^-01^ | 2.45 | 2.43 | 2.18 | | 0.25 |
| **Dozing (Sometimes vs Never/rarely)** | | | | | | | | | | |
| Average sweet liking (multi-item mean) | Standard covariates | 51659 | 0.169 | 0.013 | 1.11x10^-41^* | 3.38 | 3.35 | 2.94 | | 0.41 |
|  | + 5 sleep traits | 51659 | 0.165 | 0.013 | 1.89x10^-39^* | 3.72 | 3.69 | 2.94 | | 0.75 |
| Single item sweet food liking | Standard covariates | 51620 | 0.276 | 0.025 | 7.03x10^-29^* | 2.33 | 2.31 | 1.99 | | 0.32 |
|  | + 5 sleep traits | 51620 | 0.270 | 0.025 | 1.66x10^-27^* | 2.54 | 2.51 | 1.99 | | 0.51 |
| Total sugar (g/day) | Standard covariates | 60429 | 3.768 | 0.505 | 8.67x10^-14^* | 2.14 | 2.12 | 1.99 | | 0.13 |
|  | + 5 sleep traits | 60429 | 3.893 | 0.506 | 1.52x10^-14^* | 2.18 | 2.16 | 1.99 | | 0.17 |
| Total sugar (g/1000kcal) | Standard covariates | 60423 | 0.275 | 0.197 | 1.63x10^-01^ | 2.43 | 2.41 | 2.41 | | 0.00 |
|  | + 5 sleep traits | 60423 | 0.407 | 0.197 | 3.93x10^-02^ | 2.94 | 2.91 | 2.41 | | 0.50 |
| Free sugar (g/day) | Standard covariates | 60429 | 2.974 | 0.362 | 2.04x10^-16^* | 5.71 | 5.69 | 5.52 | | 0.17 |
|  | + 5 sleep traits | 60429 | 2.812 | 0.362 | 8.24x10^-15^* | 6.05 | 6.03 | 5.52 | | 0.51 |
| Free sugar (g/1000kcal) | Standard covariates | 60423 | 0.811 | 0.150 | 6.01x10^-08^* | 2.26 | 2.25 | 2.18 | | 0.07 |
|  | + 5 sleep traits | 60423 | 0.762 | 0.150 | 3.71x10^-07^* | 2.45 | 2.43 | 2.18 | | 0.25 |
| **Dozing (Often vs Never/rarely)** | | | | | | | | | | |
| Average sweet liking (multi-item mean) | Standard covariates | 51659 | 0.228 | 0.031 | 1.01x10^-13^* | 3.38 | 3.35 | 2.94 | | 0.41 |
|  | + 5 sleep traits | 51659 | 0.219 | 0.031 | 1.07x10^-12^* | 3.72 | 3.69 | 2.94 | | 0.75 |
| Single item sweet food liking | Standard covariates | 51620 | 0.468 | 0.061 | 1.33x10^-14^* | 2.33 | 2.31 | 1.99 | | 0.32 |
|  | + 5 sleep traits | 51620 | 0.450 | 0.061 | 1.36x10^-13^* | 2.54 | 2.51 | 1.99 | | 0.51 |
| Total sugar (g/day) | Standard covariates | 60429 | 7.268 | 1.194 | 1.16x10^-09^* | 2.14 | 2.12 | 1.99 | | 0.13 |
|  | + 5 sleep traits | 60429 | 7.661 | 1.199 | 1.70x10^-10^* | 2.18 | 2.16 | 1.99 | | 0.17 |
| Total sugar (g/1000kcal) | Standard covariates | 60423 | 0.380 | 0.466 | 4.15x10^-01^ | 2.43 | 2.41 | 2.41 | | 0.00 |
|  | + 5 sleep traits | 60423 | 0.758 | 0.467 | 1.05x10^-01^ | 2.94 | 2.91 | 2.41 | | 0.50 |
| Free sugar (g/day) | Standard covariates | 60429 | 6.307 | 0.855 | 1.66x10^-13^* | 5.71 | 5.69 | 5.52 | | 0.17 |
|  | + 5 sleep traits | 60429 | 5.874 | 0.858 | 7.50x10^-12^* | 6.05 | 6.03 | 5.52 | | 0.51 |
| Free sugar (g/1000kcal) | Standard covariates | 60423 | 1.553 | 0.354 | 1.14x10^-05^* | 2.26 | 2.25 | 2.18 | | 0.07 |
|  | + 5 sleep traits | 60423 | 1.422 | 0.355 | 6.22x10^-05^* | 2.45 | 2.43 | 2.18 | | 0.25 |
| **Composite sleep score (per unit)** | | | | | | | | | | |
| Average sweet liking (multi-item mean) | Standard covariates | 51659 | -0.069 | 0.005 | 1.38x10^-48^* | 3.36 | 3.34 | 2.94 | | 0.40 |
| Liking for sweet foods | Standard covariates | 51620 | -0.105 | 0.009 | 4.14x10^-29^* | 2.25 | 2.23 | 1.99 | | 0.24 |
| Total sugar (g/day) | Standard covariates | 60429 | -0.420 | 0.193 | 2.98x10^-02^ | 2.01 | 1.99 | 1.99 | | 0.01 |
| Total sugar (g/1000kcal) | Standard covariates | 60423 | 0.633 | 0.075 | 5.24x10^-17^* | 2.54 | 2.52 | 2.41 | | 0.11 |
| Free sugar (g/day) | Standard covariates | 60429 | -2.193 | 0.138 | 1.60x10^-56^* | 5.92 | 5.91 | 5.52 | | 0.39 |
| Free sugar (g/1000kcal) | Standard covariates | 60423 | -0.691 | 0.057 | 1.39x10^-33^* | 2.43 | 2.41 | 2.18 | | 0.23 |
| **Sensitivity Analyses (average sweet liking, complete case)** | | | | | | | | | | |
| **Chronotype (Evening vs Morning)** | | | | | | | | | | |
| Average sweet liking (multi-item mean) | Standard covariates | 45681 | 0.044 | 0.004 | 1.71x10^-29^* | 3.51 | 3.49 | | 3.23 | 0.27 |
|  | + 5 sleep traits | 45681 | 0.043 | 0.004 | 7.38x10^-29^* | 4.03 | 4.00 | | 3.23 | 0.77 |
| **Sleep duration (per hour)** | | | | | | | | | | |
| Average sweet liking (multi-item mean) | Standard covariates | 45681 | 0.009 | 0.005 | 8.69x10^-02^ | 3.25 | 3.23 | | 3.23 | 0.00 |
|  | + 5 sleep traits | 45681 | 0.013 | 0.005 | 1.59x10^-02^ | 4.03 | 4.00 | | 3.23 | 0.77 |
| **Insomnia (Sometimes vs Never/rarely)** | | | | | | | | | | |
| Average sweet liking (multi-item mean) | Standard covariates | 45681 | 0.053 | 0.013 | 2.27x10^-05^* | 3.29 | 3.26 | | 3.23 | 0.04 |
|  | Standard covariates + 5 sleep traits | 45681 | 0.053 | 0.013 | 2.57x10^-05^* | 4.03 | 4.00 | | 3.23 | 0.77 |
| **Insomnia (Usually vs Never/rarely)** | | | | | | | | | | |
| Average sweet liking (multi-item mean) | Standard covariates | 45681 | 0.046 | 0.014 | 1.21x10^-03^* | 3.29 | 3.26 | | 3.23 | 0.04 |
|  | Standard covariates + 5 sleep traits | 45681 | 0.041 | 0.015 | 4.32x10^-03^ | 4.03 | 4.00 | | 3.23 | 0.77 |
| **Snoring (Yes vs No)** | | | | | | | | | | |
| Average sweet liking (multi-item mean) | Standard covariates | 45681 | 0.067 | 0.011 | 3.34x10^-09^* | 3.32 | 3.30 | | 3.23 | 0.07 |
|  | + 5 sleep traits | 45681 | 0.056 | 0.011 | 6.40x10^-07^* | 4.03 | 4.00 | | 3.23 | 0.77 |
| **Dozing (Sometimes vs Never/rarely)** | | | | | | | | | | |
| Average sweet liking (multi-item mean) | Standard covariates | 45681 | 0.171 | 0.013 | 1.10x10^-37^* | 3.67 | 3.65 | | 3.23 | 0.42 |
|  | + 5 sleep traits | 45681 | 0.166 | 0.013 | 2.12x10^-35^* | 4.03 | 4.00 | | 3.23 | 0.77 |
| **Dozing (Often vs Never/rarely)** | | | | | | | | | | |
| Average sweet liking (multi-item mean) | Standard covariates | 45681 | 0.232 | 0.033 | 2.75x10^-12^* | 3.67 | 3.65 | | 3.23 | 0.42 |
|  | + 5 sleep traits | 45681 | 0.222 | 0.033 | 2.19x10^-11^* | 4.03 | 4.00 | | 3.23 | 0.77 |
| **Composite sleep score (per unit)** | | | | | | | | | | |
| Average sweet liking (multi-item mean) | Standard covariates | 45681 | -0.072 | 0.005 | 2.86x10^-47^* | 3.69 | 3.67 | | 3.23 | 0.44 |

Standard covariates include sex, age, body mass index, socioeconomic status (Townsend Deprivation Index), and ethnicity.

+ 5 sleep traits include chronotype, sleep duration, insomnia, snoring, daytime dozing. Composite sleep score was modeled separately.
Adjusted P-values obtained from Bonferroni-correction for multiple comparisons of 20 independent tests (see Methods).
*p < 0.0025 (Bonferroni-corrected threshold for 20 independent tests)
Columns: Adj_r^2^, adjusted R^2^ of the full regression model; Cov_adj_r^2^, adjusted R^2^ of the covariate-only model.

**Supplementary Table 4.** Results from mediation analysis.

| **Liking mediator (M)** | **Intake outcome (Y)** | **Sample size** | **Path type** | **Covariates** | **β** | **SE** | **P-value** |
| --- | --- | --- | --- | --- | --- | --- | --- |
| **Sleep predictor (X): Chronotype (Evening vs Morning)** | | | | | | | |
| Average sweet liking (multi-item mean) | Free sugar (g/1000kcal) | 35359 | a (X→M) | Standard covariates | 0.042 | 0.004 | 0.00E+00* |
|  |  |  |  | + 5 sleep traits | 0.041 | 0.004 | 0.00E+00* |
|  |  |  | b (M→Y) | Standard covariates | 2.626 | 0.063 | 0.00E+00* |
|  |  |  |  | + 5 sleep traits | 2.620 | 0.063 | 0.00E+00* |
|  |  |  | c' (X→Y) | Standard covariates | 0.357 | 0.052 | 8.05E-12* |
|  |  |  |  | + 5 sleep traits | 0.358 | 0.052 | 7.02E-12* |
|  |  |  | indirect | Standard covariates | 0.110 | 0.012 | 0.00E+00* |
|  |  |  |  | + 5 sleep traits | 0.108 | 0.012 | 0.00E+00* |
|  |  |  | total | Standard covariates | 23.5% | 0.033 | 9.17E-13* |
|  |  |  |  | + 5 sleep traits | 0.466 | 0.053 | 0.00E+00* |
|  |  |  | Proportion mediated | Standard covariates | 23.5% | 0.033 | 9.17E-13* |
|  |  |  |  | + 5 sleep traits | 23.2% | 0.033 | 1.24E-12* |
|  | Free sugar (g/day) | 35362 | a (X→M) | Standard covariates | 0.042 | 0.004 | 0.00E+00* |
|  |  |  |  | + 5 sleep traits | 0.041 | 0.004 | 0.00E+00* |
|  |  |  | b (M→Y) | Standard covariates | 6.976 | 0.153 | 0.00E+00* |
|  |  |  |  | + 5 sleep traits | 6.919 | 0.153 | 0.00E+00* |
|  |  |  | c' (X→Y) | Standard covariates | 1.100 | 0.127 | 0.00E+00* |
|  |  |  |  | + 5 sleep traits | 1.098 | 0.127 | 0.00E+00* |
|  |  |  | indirect | Standard covariates | 0.291 | 0.031 | 0.00E+00* |
|  |  |  |  | + 5 sleep traits | 0.285 | 0.031 | 0.00E+00* |
|  |  |  | total | Standard covariates | 1.391 | 0.131 | 0.00E+00* |
|  |  |  |  | + 5 sleep traits | 1.383 | 0.130 | 0.00E+00* |
|  |  |  | Proportion mediated | Standard covariates | 20.9% | 0.026 | 1.78E-15* |
|  |  |  |  | + 5 sleep traits | 20.6% | 0.026 | 3.11E-15* |
|  | Total sugar (g/1000kcal) | 35359 | a (X→M) | Standard covariates | 0.042 | 0.004 | 0.00E+00* |
|  |  |  |  | + 5 sleep traits | 0.041 | 0.004 | 0.00E+00* |
|  |  |  | b (M→Y) | Standard covariates | 1.628 | 0.086 | 0.00E+00* |
|  |  |  |  | + 5 sleep traits | 1.653 | 0.086 | 0.00E+00* |
|  |  |  | c' (X→Y) | Standard covariates | -0.772 | 0.071 | 0.00E+00* |
|  |  |  |  | + 5 sleep traits | -0.768 | 0.071 | 0.00E+00* |
|  |  |  | indirect | Standard covariates | 0.068 | 0.008 | 0.00E+00* |
|  |  |  |  | + 5 sleep traits | 0.068 | 0.008 | 0.00E+00* |
|  |  |  | total | Standard covariates | -0.704 | 0.072 | 0.00E+00* |
|  |  |  |  | + 5 sleep traits | -0.700 | 0.071 | 0.00E+00* |
|  |  |  | Proportion mediated | Standard covariates | -9.6% | 0.016 | 7.66E-10* |
|  |  |  |  | + 5 sleep traits | -9.7% | 0.016 | 9.66E-10* |
|  | Total sugar (g/day) | 35362 | a (X→M) | Standard covariates | 0.042 | 0.004 | 0.00E+00* |
|  |  |  |  | + 5 sleep traits | 0.041 | 0.004 | 0.00E+00* |
|  |  |  | b (M→Y) | Standard covariates | 6.641 | 0.218 | 0.00E+00* |
|  |  |  |  | + 5 sleep traits | 6.591 | 0.219 | 0.00E+00* |
|  |  |  | c' (X→Y) | Standard covariates | -0.807 | 0.181 | 8.61E-06* |
|  |  |  |  | + 5 sleep traits | -0.809 | 0.181 | 8.15E-06* |
|  |  |  | indirect | Standard covariates | 0.277 | 0.031 | 0.00E+00* |
|  |  |  |  | + 5 sleep traits | 0.272 | 0.030 | 0.00E+00* |
|  |  |  | total | Standard covariates | -0.530 | 0.184 | 3.87E-03 |
|  |  |  |  | + 5 sleep traits | -0.537 | 0.183 | 3.39E-03 |
|  |  |  | Proportion mediated | Standard covariates | -52.3% | 0.198 | 8.37E-03 |
|  |  |  |  | + 5 sleep traits | -50.5% | 0.189 | 7.65E-03 |
| Single item sweet food liking | Free sugar (g/1000kcal) | 35328 | a (X→M) | Standard covariates | 0.066 | 0.009 | 5.17E-14* |
|  |  |  |  | + 5 sleep traits | 0.065 | 0.009 | 8.57E-14* |
|  |  |  | b (M→Y) | Standard covariates | 1.092 | 0.032 | 0.00E+00* |
|  |  |  |  | + 5 sleep traits | 1.087 | 0.032 | 0.00E+00* |
|  |  |  | c' (X→Y) | Standard covariates | 0.396 | 0.053 | 5.31E-14* |
|  |  |  |  | + 5 sleep traits | 0.396 | 0.053 | 4.93E-14* |
|  |  |  | indirect | Standard covariates | 0.072 | 0.010 | 1.94E-13* |
|  |  |  |  | + 5 sleep traits | 0.071 | 0.010 | 3.13E-13* |
|  |  |  | total | Standard covariates | 0.468 | 0.053 | 0.00E+00* |
|  |  |  |  | + 5 sleep traits | 0.467 | 0.053 | 0.00E+00* |
|  |  |  | Proportion mediated | Standard covariates | 15.4% | 0.025 | 5.97E-10* |
|  |  |  |  | + 5 sleep traits | 15.2% | 0.025 | 7.64E-10* |
|  | Free sugar (g/day) | 35331 | a (X→M) | Standard covariates | 0.066 | 0.009 | 5.68E-14* |
|  |  |  |  | + 5 sleep traits | 0.065 | 0.009 | 9.39E-14* |
|  |  |  | b (M→Y) | Standard covariates | 2.862 | 0.078 | 0.00E+00* |
|  |  |  |  | + 5 sleep traits | 2.832 | 0.078 | 0.00E+00* |
|  |  |  | c' (X→Y) | Standard covariates | 1.205 | 0.128 | 0.00E+00* |
|  |  |  |  | + 5 sleep traits | 1.201 | 0.128 | 0.00E+00* |
|  |  |  | indirect | Standard covariates | 0.189 | 0.026 | 1.79E-13* |
|  |  |  |  | + 5 sleep traits | 0.185 | 0.025 | 2.91E-13* |
|  |  |  | total | Standard covariates | 1.394 | 0.131 | 0.00E+00* |
|  |  |  |  | + 5 sleep traits | 1.386 | 0.130 | 0.00E+00* |
|  |  |  | Proportion mediated | Standard covariates | 13.6% | 0.020 | 2.40E-11* |
|  |  |  |  | + 5 sleep traits | 13.3% | 0.020 | 3.61E-11* |
|  | Total sugar (g/1000kcal) | 35328 | a (X→M) | Standard covariates | 0.066 | 0.009 | 5.17E-14* |
|  |  |  |  | + 5 sleep traits | 0.065 | 0.009 | 8.57E-14* |
|  |  |  | b (M→Y) | Standard covariates | 0.742 | 0.043 | 0.00E+00* |
|  |  |  |  | + 5 sleep traits | 0.751 | 0.043 | 0.00E+00* |
|  |  |  | c' (X→Y) | Standard covariates | -0.750 | 0.071 | 0.00E+00* |
|  |  |  |  | + 5 sleep traits | -0.746 | 0.071 | 0.00E+00* |
|  |  |  | indirect | Standard covariates | 0.049 | 0.007 | 5.38E-12* |
|  |  |  |  | + 5 sleep traits | 0.049 | 0.007 | 7.02E-12* |
|  |  |  | total | Standard covariates | -0.701 | 0.072 | 0.00E+00* |
|  |  |  |  | + 5 sleep traits | -0.697 | 0.071 | 0.00E+00* |
|  |  |  | Proportion mediated | Standard covariates | -7.0% | 0.013 | 5.63E-08* |
|  |  |  |  | + 5 sleep traits | -7.1% | 0.013 | 6.72E-08* |
|  | Total sugar (g/day) | 35331 | a (X→M) | Standard covariates | 0.066 | 0.009 | 5.68E-14* |
|  |  |  |  | + 5 sleep traits | 0.065 | 0.009 | 9.39E-14* |
|  |  |  | b (M→Y) | Standard covariates | 2.785 | 0.110 | 0.00E+00* |
|  |  |  |  | + 5 sleep traits | 2.759 | 0.110 | 0.00E+00* |
|  |  |  | c' (X→Y) | Standard covariates | -0.707 | 0.182 | 1.03E-04* |
|  |  |  |  | + 5 sleep traits | -0.710 | 0.182 | 9.55E-05* |
|  |  |  | indirect | Standard covariates | 0.184 | 0.026 | 5.87E-13* |
|  |  |  |  | + 5 sleep traits | 0.180 | 0.025 | 9.40E-13* |
|  |  |  | total | Standard covariates | -0.523 | 0.184 | 4.37E-03 |
|  |  |  |  | + 5 sleep traits | -0.530 | 0.183 | 3.87E-03 |
|  |  |  | Proportion mediated | Standard covariates | -35.1% | 0.138 | 1.10E-02 |
|  |  |  |  | + 5 sleep traits | -34.0% | 0.132 | 1.02E-02 |
| **Sleep predictor (X): Sleep duration (per hour)** | | | | | | | |
| Average sweet liking (multi-item mean) | Free sugar (g/1000kcal) | 35359 | a (X→M) | Standard covariates | 0.012 | 0.006 | 4.39E-02 |
|  |  |  |  | + 5 sleep traits | 0.017 | 0.006 | 4.83E-03 |
|  |  |  | b (M→Y) | Standard covariates | 2.649 | 0.063 | 0.00E+00* |
|  |  |  |  | + 5 sleep traits | 2.620 | 0.063 | 0.00E+00* |
|  |  |  | c' (X→Y) | Standard covariates | -0.170 | 0.069 | 1.40E-02 |
|  |  |  |  | + 5 sleep traits | -0.138 | 0.071 | 5.34E-02 |
|  |  |  | indirect | Standard covariates | 0.031 | 0.016 | 4.42E-02 |
|  |  |  |  | + 5 sleep traits | 0.044 | 0.016 | 4.93E-03 |
|  |  |  | total | Standard covariates | -0.139 | 0.071 | 5.04E-02 |
|  |  |  |  | + 5 sleep traits | -0.093 | 0.073 | 2.02E-01 |
|  |  |  | Proportion mediated | Standard covariates | -22.6% | 0.178 | 2.04E-01 |
|  |  |  |  | + 5 sleep traits | -47.7% | 0.443 | 2.81E-01 |
|  | Free sugar (g/day) | 35362 | a (X→M) | Standard covariates | 0.012 | 0.006 | 4.33E-02 |
|  |  |  |  | + 5 sleep traits | 0.017 | 0.006 | 4.71E-03 |
|  |  |  | b (M→Y) | Standard covariates | 7.045 | 0.153 | 0.00E+00* |
|  |  |  |  | + 5 sleep traits | 6.919 | 0.153 | 0.00E+00* |
|  |  |  | c' (X→Y) | Standard covariates | -0.304 | 0.168 | 7.04E-02 |
|  |  |  |  | + 5 sleep traits | -0.207 | 0.173 | 2.33E-01 |
|  |  |  | indirect | Standard covariates | 0.083 | 0.041 | 4.35E-02 |
|  |  |  |  | + 5 sleep traits | 0.118 | 0.042 | 4.79E-03 |
|  |  |  | total | Standard covariates | -0.221 | 0.173 | 2.02E-01 |
|  |  |  |  | + 5 sleep traits | -0.089 | 0.178 | 6.17E-01 |
|  |  |  | Proportion mediated | Standard covariates | -37.7% | 0.385 | 3.28E-01 |
|  |  |  |  | + 5 sleep traits | -132.1% | 2.787 | 6.36E-01 |
|  | Total sugar (g/1000kcal) | 35359 | a (X→M) | Standard covariates | 0.012 | 0.006 | 4.39E-02 |
|  |  |  |  | + 5 sleep traits | 0.017 | 0.006 | 4.83E-03 |
|  |  |  | b (M→Y) | Standard covariates | 1.585 | 0.086 | 0.00E+00* |
|  |  |  |  | + 5 sleep traits | 1.653 | 0.086 | 0.00E+00* |
|  |  |  | c' (X→Y) | Standard covariates | -0.337 | 0.094 | 3.53E-04* |
|  |  |  |  | + 5 sleep traits | -0.412 | 0.097 | 2.23E-05* |
|  |  |  | indirect | Standard covariates | 0.019 | 0.009 | 4.52E-02 |
|  |  |  |  | + 5 sleep traits | 0.028 | 0.010 | 5.29E-03 |
|  |  |  | total | Standard covariates | -0.319 | 0.095 | 7.83E-04* |
|  |  |  |  | + 5 sleep traits | -0.384 | 0.098 | 8.45E-05* |
|  |  |  | Proportion mediated | Standard covariates | -5.9% | 0.036 | 9.87E-02 |
|  |  |  |  | + 5 sleep traits | -7.3% | 0.034 | 2.97E-02 |
|  | Total sugar (g/day) | 35362 | a (X→M) | Standard covariates | 0.012 | 0.006 | 4.33E-02 |
|  |  |  |  | + 5 sleep traits | 0.017 | 0.006 | 4.71E-03 |
|  |  |  | b (M→Y) | Standard covariates | 6.598 | 0.218 | 0.00E+00* |
|  |  |  |  | + 5 sleep traits | 6.591 | 0.219 | 0.00E+00* |
|  |  |  | c' (X→Y) | Standard covariates | -0.506 | 0.240 | 3.52E-02 |
|  |  |  |  | + 5 sleep traits | -0.588 | 0.247 | 1.74E-02 |
|  |  |  | indirect | Standard covariates | 0.078 | 0.039 | 4.38E-02 |
|  |  |  |  | + 5 sleep traits | 0.112 | 0.040 | 4.90E-03 |
|  |  |  | total | Standard covariates | -0.428 | 0.243 | 7.86E-02 |
|  |  |  |  | + 5 sleep traits | -0.476 | 0.250 | 5.74E-02 |
|  |  |  | Proportion mediated | Standard covariates | -18.2% | 0.148 | 2.18E-01 |
|  |  |  |  | + 5 sleep traits | -23.5% | 0.160 | 1.41E-01 |
| Single item sweet food liking | Free sugar (g/1000kcal) | 35328 | a (X→M) | Standard covariates | 0.018 | 0.012 | 1.24E-01 |
|  |  |  |  | + 5 sleep traits | 0.033 | 0.012 | 5.60E-03 |
|  |  |  | b (M→Y) | Standard covariates | 1.102 | 0.032 | 0.00E+00* |
|  |  |  |  | + 5 sleep traits | 1.087 | 0.032 | 0.00E+00* |
|  |  |  | c' (X→Y) | Standard covariates | -0.156 | 0.070 | 2.50E-02 |
|  |  |  |  | + 5 sleep traits | -0.127 | 0.072 | 7.63E-02 |
|  |  |  | indirect | Standard covariates | 0.020 | 0.013 | 1.24E-01 |
|  |  |  |  | + 5 sleep traits | 0.036 | 0.013 | 5.75E-03 |
|  |  |  | total | Standard covariates | -0.136 | 0.071 | 5.42E-02 |
|  |  |  |  | + 5 sleep traits | -0.091 | 0.073 | 2.11E-01 |
|  |  |  | Proportion mediated | Standard covariates | -14.5% | 0.131 | 2.68E-01 |
|  |  |  |  | + 5 sleep traits | -39.6% | 0.370 | 2.85E-01 |
|  | Free sugar (g/day) | 35331 | a (X→M) | Standard covariates | 0.018 | 0.012 | 1.20E-01 |
|  |  |  |  | + 5 sleep traits | 0.033 | 0.012 | 5.31E-03 |
|  |  |  | b (M→Y) | Standard covariates | 2.892 | 0.078 | 0.00E+00* |
|  |  |  |  | + 5 sleep traits | 2.832 | 0.078 | 0.00E+00* |
|  |  |  | c' (X→Y) | Standard covariates | -0.270 | 0.170 | 1.12E-01 |
|  |  |  |  | + 5 sleep traits | -0.180 | 0.175 | 3.04E-01 |
|  |  |  | indirect | Standard covariates | 0.052 | 0.034 | 1.20E-01 |
|  |  |  |  | + 5 sleep traits | 0.095 | 0.034 | 5.45E-03 |
|  |  |  | total | Standard covariates | -0.218 | 0.173 | 2.09E-01 |
|  |  |  |  | + 5 sleep traits | -0.085 | 0.178 | 6.32E-01 |
|  |  |  | Proportion mediated | Standard covariates | -24.1% | 0.269 | 3.71E-01 |
|  |  |  |  | + 5 sleep traits | -110.9% | 2.426 | 6.48E-01 |
|  | Total sugar (g/1000kcal) | 35328 | a (X→M) | Standard covariates | 0.018 | 0.012 | 1.24E-01 |
|  |  |  |  | + 5 sleep traits | 0.033 | 0.012 | 5.60E-03 |
|  |  |  | b (M→Y) | Standard covariates | 0.725 | 0.043 | 0.00E+00* |
|  |  |  |  | + 5 sleep traits | 0.751 | 0.043 | 0.00E+00* |
|  |  |  | c' (X→Y) | Standard covariates | -0.330 | 0.095 | 4.81E-04* |
|  |  |  |  | + 5 sleep traits | -0.406 | 0.097 | 2.91E-05* |
|  |  |  | indirect | Standard covariates | 0.013 | 0.008 | 1.25E-01 |
|  |  |  |  | + 5 sleep traits | 0.025 | 0.009 | 6.22E-03 |
|  |  |  | total | Standard covariates | -0.317 | 0.095 | 8.37E-04* |
|  |  |  |  | + 5 sleep traits | -0.381 | 0.098 | 9.33E-05* |
|  |  |  | Proportion mediated | Standard covariates | -4.1% | 0.030 | 1.77E-01 |
|  |  |  |  | + 5 sleep traits | -6.5% | 0.030 | 3.15E-02 |
|  | Total sugar (g/day) | 35331 | a (X→M) | Standard covariates | 0.018 | 0.012 | 1.20E-01 |
|  |  |  |  | + 5 sleep traits | 0.033 | 0.012 | 5.31E-03 |
|  |  |  | b (M→Y) | Standard covariates | 2.770 | 0.110 | 0.00E+00* |
|  |  |  |  | + 5 sleep traits | 2.759 | 0.110 | 0.00E+00* |
|  |  |  | c' (X→Y) | Standard covariates | -0.475 | 0.241 | 4.91E-02 |
|  |  |  |  | + 5 sleep traits | -0.562 | 0.248 | 2.36E-02 |
|  |  |  | indirect | Standard covariates | 0.050 | 0.032 | 1.21E-01 |
|  |  |  |  | + 5 sleep traits | 0.092 | 0.033 | 5.60E-03 |
|  |  |  | total | Standard covariates | -0.425 | 0.243 | 8.11E-02 |
|  |  |  |  | + 5 sleep traits | -0.470 | 0.251 | 6.07E-02 |
|  |  |  | Proportion mediated | Standard covariates | -11.8% | 0.108 | 2.76E-01 |
|  |  |  |  | + 5 sleep traits | -19.6% | 0.134 | 1.42E-01 |
| **Sleep predictor (X): Insomnia (Usually vs Sometimes vs Never/rarely; treated as continuous)** | | | | | | | |
| Average sweet liking (multi-item mean) | Free sugar (g/1000kcal) | 35359 | a (X→M) | Standard covariates | 0.020 | 0.008 | 1.32E-02 |
|  |  |  |  | + 5 sleep traits | 0.020 | 0.008 | 1.66E-02 |
|  |  |  | b (M→Y) | Standard covariates | 2.646 | 0.063 | 0.00E+00* |
|  |  |  |  | + 5 sleep traits | 2.620 | 0.063 | 0.00E+00* |
|  |  |  | c' (X→Y) | Standard covariates | 0.226 | 0.094 | 1.66E-02 |
|  |  |  |  | + 5 sleep traits | 0.177 | 0.097 | 6.89E-02 |
|  |  |  | indirect | Standard covariates | 0.052 | 0.021 | 1.33E-02 |
|  |  |  |  | + 5 sleep traits | 0.052 | 0.022 | 1.68E-02 |
|  |  |  | total | Standard covariates | 0.279 | 0.097 | 3.97E-03 |
|  |  |  |  | + 5 sleep traits | 0.229 | 0.100 | 2.18E-02 |
|  |  |  | Proportion mediated | Standard covariates | 18.8% | 0.089 | 3.40E-02 |
|  |  |  |  | + 5 sleep traits | 22.6% | 0.121 | 6.16E-02 |
|  | Free sugar (g/day) | 35362 | a (X→M) | Standard covariates | 0.020 | 0.008 | 1.31E-02 |
|  |  |  |  | + 5 sleep traits | 0.020 | 0.008 | 1.64E-02 |
|  |  |  | b (M→Y) | Standard covariates | 7.037 | 0.153 | 0.00E+00* |
|  |  |  |  | + 5 sleep traits | 6.919 | 0.153 | 0.00E+00* |
|  |  |  | c' (X→Y) | Standard covariates | 0.592 | 0.230 | 1.00E-02 |
|  |  |  |  | + 5 sleep traits | 0.466 | 0.237 | 4.92E-02 |
|  |  |  | indirect | Standard covariates | 0.140 | 0.056 | 1.32E-02 |
|  |  |  |  | + 5 sleep traits | 0.137 | 0.057 | 1.65E-02 |
|  |  |  | total | Standard covariates | 0.732 | 0.237 | 1.98E-03* |
|  |  |  |  | + 5 sleep traits | 0.603 | 0.244 | 1.34E-02 |
|  |  |  | Proportion mediated | Standard covariates | 19.1% | 0.087 | 2.74E-02 |
|  |  |  |  | + 5 sleep traits | 22.7% | 0.115 | 4.93E-02 |
|  | Total sugar (g/1000kcal) | 35359 | a (X→M) | Standard covariates | 0.020 | 0.008 | 1.32E-02 |
|  |  |  |  | + 5 sleep traits | 0.020 | 0.008 | 1.66E-02 |
|  |  |  | b (M→Y) | Standard covariates | 1.585 | 0.086 | 0.00E+00* |
|  |  |  |  | + 5 sleep traits | 1.653 | 0.086 | 0.00E+00* |
|  |  |  | c' (X→Y) | Standard covariates | -0.404 | 0.129 | 1.73E-03* |
|  |  |  |  | + 5 sleep traits | -0.569 | 0.133 | 1.81E-05* |
|  |  |  | indirect | Standard covariates | 0.031 | 0.013 | 1.40E-02 |
|  |  |  |  | + 5 sleep traits | 0.033 | 0.014 | 1.75E-02 |
|  |  |  | total | Standard covariates | -0.373 | 0.130 | 4.03E-03 |
|  |  |  |  | + 5 sleep traits | -0.537 | 0.134 | 5.79E-05* |
|  |  |  | Proportion mediated | Standard covariates | -8.4% | 0.047 | 7.44E-02 |
|  |  |  |  | + 5 sleep traits | -6.1% | 0.031 | 4.99E-02 |
|  | Total sugar (g/day) | 35362 | a (X→M) | Standard covariates | 0.020 | 0.008 | 1.31E-02 |
|  |  |  |  | + 5 sleep traits | 0.020 | 0.008 | 1.64E-02 |
|  |  |  | b (M→Y) | Standard covariates | 6.598 | 0.218 | 0.00E+00* |
|  |  |  |  | + 5 sleep traits | 6.591 | 0.219 | 0.00E+00* |
|  |  |  | c' (X→Y) | Standard covariates | -0.564 | 0.328 | 8.55E-02 |
|  |  |  |  | + 5 sleep traits | -0.928 | 0.338 | 6.11E-03 |
|  |  |  | indirect | Standard covariates | 0.131 | 0.053 | 1.34E-02 |
|  |  |  |  | + 5 sleep traits | 0.130 | 0.054 | 1.67E-02 |
|  |  |  | total | Standard covariates | -0.433 | 0.332 | 1.92E-01 |
|  |  |  |  | + 5 sleep traits | -0.798 | 0.343 | 1.99E-02 |
|  |  |  | Proportion mediated | Standard covariates | -30.2% | 0.279 | 2.78E-01 |
|  |  |  |  | + 5 sleep traits | -16.3% | 0.105 | 1.21E-01 |
| Single item sweet food liking | Free sugar (g/1000kcal) | 35328 | a (X→M) | Standard covariates | 0.067 | 0.016 | 2.62E-05* |
|  |  |  |  | + 5 sleep traits | 0.067 | 0.016 | 3.99E-05* |
|  |  |  | b (M→Y) | Standard covariates | 1.100 | 0.032 | 0.00E+00* |
|  |  |  |  | + 5 sleep traits | 1.087 | 0.032 | 0.00E+00* |
|  |  |  | c' (X→Y) | Standard covariates | 0.204 | 0.095 | 3.23E-02 |
|  |  |  |  | + 5 sleep traits | 0.155 | 0.098 | 1.15E-01 |
|  |  |  | indirect | Standard covariates | 0.074 | 0.018 | 3.00E-05* |
|  |  |  |  | + 5 sleep traits | 0.073 | 0.018 | 4.53E-05* |
|  |  |  | total | Standard covariates | 0.277 | 0.097 | 4.16E-03 |
|  |  |  |  | + 5 sleep traits | 0.228 | 0.100 | 2.25E-02 |
|  |  |  | Proportion mediated | Standard covariates | 26.5% | 0.102 | 9.63E-03 |
|  |  |  |  | + 5 sleep traits | 32.1% | 0.149 | 3.07E-02 |
|  | Free sugar (g/day) | 35331 | a (X→M) | Standard covariates | 0.067 | 0.016 | 2.57E-05* |
|  |  |  |  | + 5 sleep traits | 0.067 | 0.016 | 3.85E-05* |
|  |  |  | b (M→Y) | Standard covariates | 2.887 | 0.078 | 0.00E+00* |
|  |  |  |  | + 5 sleep traits | 2.832 | 0.078 | 0.00E+00* |
|  |  |  | c' (X→Y) | Standard covariates | 0.541 | 0.232 | 2.00E-02 |
|  |  |  |  | + 5 sleep traits | 0.414 | 0.239 | 8.40E-02 |
|  |  |  | indirect | Standard covariates | 0.193 | 0.046 | 2.89E-05* |
|  |  |  |  | + 5 sleep traits | 0.191 | 0.047 | 4.31E-05* |
|  |  |  | total | Standard covariates | 0.734 | 0.237 | 1.94E-03* |
|  |  |  |  | + 5 sleep traits | 0.605 | 0.244 | 1.31E-02 |
|  |  |  | Proportion mediated | Standard covariates | 26.3% | 0.096 | 5.82E-03 |
|  |  |  |  | + 5 sleep traits | 31.6% | 0.136 | 2.01E-02 |
|  | Total sugar (g/1000kcal) | 35328 | a (X→M) | Standard covariates | 0.067 | 0.016 | 2.62E-05* |
|  |  |  |  | + 5 sleep traits | 0.067 | 0.016 | 3.99E-05* |
|  |  |  | b (M→Y) | Standard covariates | 0.727 | 0.043 | 0.00E+00* |
|  |  |  |  | + 5 sleep traits | 0.751 | 0.043 | 0.00E+00* |
|  |  |  | c' (X→Y) | Standard covariates | -0.420 | 0.129 | 1.17E-03* |
|  |  |  |  | + 5 sleep traits | -0.585 | 0.133 | 1.12E-05* |
|  |  |  | indirect | Standard covariates | 0.049 | 0.012 | 4.53E-05* |
|  |  |  |  | + 5 sleep traits | 0.051 | 0.013 | 6.38E-05* |
|  |  |  | total | Standard covariates | -0.371 | 0.130 | 4.24E-03 |
|  |  |  |  | + 5 sleep traits | -0.534 | 0.134 | 6.42E-05* |
|  |  |  | Proportion mediated | Standard covariates | -13.1% | 0.058 | 2.43E-02 |
|  |  |  |  | + 5 sleep traits | -9.5% | 0.035 | 6.77E-03 |
|  | Total sugar (g/day) | 35331 | a (X→M) | Standard covariates | 0.067 | 0.016 | 2.57E-05* |
|  |  |  |  | + 5 sleep traits | 0.067 | 0.016 | 3.85E-05* |
|  |  |  | b (M→Y) | Standard covariates | 2.773 | 0.110 | 0.00E+00* |
|  |  |  |  | + 5 sleep traits | 2.759 | 0.110 | 0.00E+00* |
|  |  |  | c' (X→Y) | Standard covariates | -0.609 | 0.330 | 6.48E-02 |
|  |  |  |  | + 5 sleep traits | -0.973 | 0.340 | 4.23E-03 |
|  |  |  | indirect | Standard covariates | 0.186 | 0.045 | 3.31E-05* |
|  |  |  |  | + 5 sleep traits | 0.186 | 0.046 | 4.87E-05* |
|  |  |  | total | Standard covariates | -0.423 | 0.333 | 2.03E-01 |
|  |  |  |  | + 5 sleep traits | -0.787 | 0.343 | 2.18E-02 |
|  |  |  | Proportion mediated | Standard covariates | -43.9% | 0.373 | 2.40E-01 |
|  |  |  |  | + 5 sleep traits | -23.7% | 0.125 | 5.82E-02 |
| **Sleep predictor (X): Snoring (Yes vs No)** | | | | | | | |
| Average sweet liking (multi-item mean) | Free sugar (g/1000kcal) | 35359 | a (X→M) | Standard covariates | 0.057 | 0.013 | 9.17E-06* |
|  |  |  |  | + 5 sleep traits | 0.046 | 0.013 | 3.35E-04* |
|  |  |  | b (M→Y) | Standard covariates | 2.648 | 0.063 | 0.00E+00* |
|  |  |  |  | + 5 sleep traits | 2.620 | 0.063 | 0.00E+00* |
|  |  |  | c' (X→Y) | Standard covariates | 0.014 | 0.152 | 9.24E-01 |
|  |  |  |  | + 5 sleep traits | 0.004 | 0.152 | 9.81E-01 |
|  |  |  | indirect | Standard covariates | 0.151 | 0.034 | 1.03E-05* |
|  |  |  |  | + 5 sleep traits | 0.121 | 0.034 | 3.52E-04* |
|  |  |  | total | Standard covariates | 0.166 | 0.156 | 2.88E-01 |
|  |  |  |  | + 5 sleep traits | 0.125 | 0.156 | 4.24E-01 |
|  |  |  | Proportion mediated | Standard covariates | 91.3% | 0.838 | 2.76E-01 |
|  |  |  |  | + 5 sleep traits | 97.1% | 1.187 | 4.13E-01 |
|  | Free sugar (g/day) | 35362 | a (X→M) | Standard covariates | 0.057 | 0.013 | 9.11E-06* |
|  |  |  |  | + 5 sleep traits | 0.046 | 0.013 | 3.32E-04* |
|  |  |  | b (M→Y) | Standard covariates | 7.033 | 0.153 | 0.00E+00* |
|  |  |  |  | + 5 sleep traits | 6.919 | 0.153 | 0.00E+00* |
|  |  |  | c' (X→Y) | Standard covariates | 0.897 | 0.370 | 1.53E-02 |
|  |  |  |  | + 5 sleep traits | 0.779 | 0.370 | 3.54E-02 |
|  |  |  | indirect | Standard covariates | 0.402 | 0.091 | 1.00E-05* |
|  |  |  |  | + 5 sleep traits | 0.319 | 0.089 | 3.47E-04* |
|  |  |  | total | Standard covariates | 1.299 | 0.381 | 6.44E-04* |
|  |  |  |  | + 5 sleep traits | 1.098 | 0.381 | 3.92E-03 |
|  |  |  | Proportion mediated | Standard covariates | 30.9% | 0.101 | 2.10E-03* |
|  |  |  |  | + 5 sleep traits | 29.1% | 0.114 | 1.06E-02 |
|  | Total sugar (g/1000kcal) | 35359 | a (X→M) | Standard covariates | 0.057 | 0.013 | 9.17E-06* |
|  |  |  |  | + 5 sleep traits | 0.046 | 0.013 | 3.35E-04* |
|  |  |  | b (M→Y) | Standard covariates | 1.600 | 0.086 | 0.00E+00* |
|  |  |  |  | + 5 sleep traits | 1.653 | 0.086 | 0.00E+00* |
|  |  |  | c' (X→Y) | Standard covariates | -1.946 | 0.207 | 0.00E+00* |
|  |  |  |  | + 5 sleep traits | -1.925 | 0.208 | 0.00E+00* |
|  |  |  | indirect | Standard covariates | 0.091 | 0.021 | 1.59E-05* |
|  |  |  |  | + 5 sleep traits | 0.076 | 0.022 | 4.21E-04* |
|  |  |  | total | Standard covariates | -1.855 | 0.208 | 0.00E+00* |
|  |  |  |  | + 5 sleep traits | -1.849 | 0.209 | 0.00E+00* |
|  |  |  | Proportion mediated | Standard covariates | -4.9% | 0.013 | 1.81E-04* |
|  |  |  |  | + 5 sleep traits | -4.1% | 0.013 | 1.53E-03* |
|  | Total sugar (g/day) | 35362 | a (X→M) | Standard covariates | 0.057 | 0.013 | 9.11E-06* |
|  |  |  |  | + 5 sleep traits | 0.046 | 0.013 | 3.32E-04* |
|  |  |  | b (M→Y) | Standard covariates | 6.611 | 0.218 | 0.00E+00* |
|  |  |  |  | + 5 sleep traits | 6.591 | 0.219 | 0.00E+00* |
|  |  |  | c' (X→Y) | Standard covariates | -1.918 | 0.528 | 2.82E-04* |
|  |  |  |  | + 5 sleep traits | -2.090 | 0.529 | 7.80E-05* |
|  |  |  | indirect | Standard covariates | 0.378 | 0.086 | 1.13E-05* |
|  |  |  |  | + 5 sleep traits | 0.304 | 0.085 | 3.65E-04* |
|  |  |  | total | Standard covariates | -1.540 | 0.535 | 3.98E-03 |
|  |  |  |  | + 5 sleep traits | -1.785 | 0.536 | 8.58E-04* |
|  |  |  | Proportion mediated | Standard covariates | -24.5% | 0.109 | 2.44E-02 |
|  |  |  |  | + 5 sleep traits | -17.0% | 0.075 | 2.36E-02 |
| Single item sweet food liking | Free sugar (g/1000kcal) | 35328 | a (X→M) | Standard covariates | 0.066 | 0.026 | 9.46E-03 |
|  |  |  |  | + 5 sleep traits | 0.048 | 0.026 | 6.30E-02 |
|  |  |  | b (M→Y) | Standard covariates | 1.101 | 0.032 | 0.00E+00* |
|  |  |  |  | + 5 sleep traits | 1.087 | 0.032 | 0.00E+00* |
|  |  |  | c' (X→Y) | Standard covariates | 0.092 | 0.153 | 5.47E-01 |
|  |  |  |  | + 5 sleep traits | 0.072 | 0.153 | 6.40E-01 |
|  |  |  | indirect | Standard covariates | 0.073 | 0.028 | 9.67E-03 |
|  |  |  |  | + 5 sleep traits | 0.052 | 0.028 | 6.34E-02 |
|  |  |  | total | Standard covariates | 0.165 | 0.156 | 2.89E-01 |
|  |  |  |  | + 5 sleep traits | 0.123 | 0.156 | 4.29E-01 |
|  |  |  | Proportion mediated | Standard covariates | 44.2% | 0.421 | 2.93E-01 |
|  |  |  |  | + 5 sleep traits | 41.9% | 0.537 | 4.35E-01 |
|  | Free sugar (g/day) | 35331 | a (X→M) | Standard covariates | 0.067 | 0.026 | 9.39E-03 |
|  |  |  |  | + 5 sleep traits | 0.048 | 0.026 | 6.26E-02 |
|  |  |  | b (M→Y) | Standard covariates | 2.888 | 0.078 | 0.00E+00* |
|  |  |  |  | + 5 sleep traits | 2.832 | 0.078 | 0.00E+00* |
|  |  |  | c' (X→Y) | Standard covariates | 1.101 | 0.374 | 3.21E-03 |
|  |  |  |  | + 5 sleep traits | 0.955 | 0.374 | 1.07E-02 |
|  |  |  | indirect | Standard covariates | 0.192 | 0.074 | 9.57E-03 |
|  |  |  |  | + 5 sleep traits | 0.135 | 0.073 | 6.30E-02 |
|  |  |  | total | Standard covariates | 1.293 | 0.381 | 6.86E-04* |
|  |  |  |  | + 5 sleep traits | 1.090 | 0.381 | 4.22E-03 |
|  |  |  | Proportion mediated | Standard covariates | 14.9% | 0.065 | 2.24E-02 |
|  |  |  |  | + 5 sleep traits | 12.4% | 0.072 | 8.63E-02 |
|  | Total sugar (g/1000kcal) | 35328 | a (X→M) | Standard covariates | 0.066 | 0.026 | 9.46E-03 |
|  |  |  |  | + 5 sleep traits | 0.048 | 0.026 | 6.30E-02 |
|  |  |  | b (M→Y) | Standard covariates | 0.729 | 0.043 | 0.00E+00* |
|  |  |  |  | + 5 sleep traits | 0.751 | 0.043 | 0.00E+00* |
|  |  |  | c' (X→Y) | Standard covariates | -1.907 | 0.208 | 0.00E+00* |
|  |  |  |  | + 5 sleep traits | -1.888 | 0.208 | 0.00E+00* |
|  |  |  | indirect | Standard covariates | 0.048 | 0.019 | 1.03E-02 |
|  |  |  |  | + 5 sleep traits | 0.036 | 0.019 | 6.45E-02 |
|  |  |  | total | Standard covariates | -1.858 | 0.209 | 0.00E+00* |
|  |  |  |  | + 5 sleep traits | -1.852 | 0.209 | 0.00E+00* |
|  |  |  | Proportion mediated | Standard covariates | -2.6% | 0.011 | 1.60E-02 |
|  |  |  |  | + 5 sleep traits | -1.9% | 0.011 | 7.55E-02 |
|  | Total sugar (g/day) | 35331 | a (X→M) | Standard covariates | 0.067 | 0.026 | 9.39E-03 |
|  |  |  |  | + 5 sleep traits | 0.048 | 0.026 | 6.26E-02 |
|  |  |  | b (M→Y) | Standard covariates | 2.773 | 0.110 | 0.00E+00* |
|  |  |  |  | + 5 sleep traits | 2.759 | 0.110 | 0.00E+00* |
|  |  |  | c' (X→Y) | Standard covariates | -1.746 | 0.530 | 9.95E-04* |
|  |  |  |  | + 5 sleep traits | -1.940 | 0.531 | 2.61E-04* |
|  |  |  | indirect | Standard covariates | 0.184 | 0.071 | 9.78E-03 |
|  |  |  |  | + 5 sleep traits | 0.132 | 0.071 | 6.34E-02 |
|  |  |  | total | Standard covariates | -1.562 | 0.535 | 3.52E-03 |
|  |  |  |  | + 5 sleep traits | -1.808 | 0.536 | 7.40E-04* |
|  |  |  | Proportion mediated | Standard covariates | -11.8% | 0.065 | 6.89E-02 |
|  |  |  |  | + 5 sleep traits | -7.3% | 0.047 | 1.23E-01 |
| **Sleep predictor (X): Dozing (Often vs Sometimes vs Never/rarely; treated as continuous)** | | | | | | | |
| Average sweet liking (multi-item mean) | Free sugar (g/1000kcal) | 35359 | a (X→M) | Standard covariates | 0.144 | 0.012 | 0.00E+00* |
|  |  |  |  | + 5 sleep traits | 0.140 | 0.012 | 0.00E+00* |
|  |  |  | b (M→Y) | Standard covariates | 2.641 | 0.063 | 0.00E+00* |
|  |  |  |  | + 5 sleep traits | 2.620 | 0.063 | 0.00E+00* |
|  |  |  | c' (X→Y) | Standard covariates | 0.226 | 0.143 | 1.14E-01 |
|  |  |  |  | + 5 sleep traits | 0.192 | 0.144 | 1.82E-01 |
|  |  |  | indirect | Standard covariates | 0.380 | 0.033 | 0.00E+00* |
|  |  |  |  | + 5 sleep traits | 0.366 | 0.033 | 0.00E+00* |
|  |  |  | total | Standard covariates | 0.607 | 0.147 | 3.47E-05* |
|  |  |  |  | + 5 sleep traits | 0.558 | 0.147 | 1.50E-04* |
|  |  |  | Proportion mediated | Standard covariates | 62.7% | 0.150 | 2.86E-05* |
|  |  |  |  | + 5 sleep traits | 65.6% | 0.171 | 1.23E-04* |
|  | Free sugar (g/day) | 35362 | a (X→M) | Standard covariates | 0.144 | 0.012 | 0.00E+00* |
|  |  |  |  | + 5 sleep traits | 0.139 | 0.012 | 0.00E+00* |
|  |  |  | b (M→Y) | Standard covariates | 6.991 | 0.153 | 0.00E+00* |
|  |  |  |  | + 5 sleep traits | 6.919 | 0.153 | 0.00E+00* |
|  |  |  | c' (X→Y) | Standard covariates | 1.833 | 0.349 | 1.47E-07* |
|  |  |  |  | + 5 sleep traits | 1.707 | 0.350 | 1.07E-06* |
|  |  |  | indirect | Standard covariates | 1.007 | 0.087 | 0.00E+00* |
|  |  |  |  | + 5 sleep traits | 0.965 | 0.087 | 0.00E+00* |
|  |  |  | total | Standard covariates | 2.839 | 0.358 | 2.22E-15* |
|  |  |  |  | + 5 sleep traits | 2.673 | 0.359 | 1.01E-13* |
|  |  |  | Proportion mediated | Standard covariates | 35.4% | 0.048 | 1.79E-13* |
|  |  |  |  | + 5 sleep traits | 36.1% | 0.052 | 3.48E-12* |
|  | Total sugar (g/1000kcal) | 35359 | a (X→M) | Standard covariates | 0.144 | 0.012 | 0.00E+00* |
|  |  |  |  | + 5 sleep traits | 0.140 | 0.012 | 0.00E+00* |
|  |  |  | b (M→Y) | Standard covariates | 1.583 | 0.086 | 0.00E+00* |
|  |  |  |  | + 5 sleep traits | 1.653 | 0.086 | 0.00E+00* |
|  |  |  | c' (X→Y) | Standard covariates | -0.058 | 0.196 | 7.67E-01 |
|  |  |  |  | + 5 sleep traits | 0.094 | 0.196 | 6.33E-01 |
|  |  |  | indirect | Standard covariates | 0.228 | 0.023 | 0.00E+00* |
|  |  |  |  | + 5 sleep traits | 0.231 | 0.023 | 0.00E+00* |
|  |  |  | total | Standard covariates | 0.170 | 0.196 | 3.87E-01 |
|  |  |  |  | + 5 sleep traits | 0.324 | 0.197 | 9.94E-02 |
|  |  |  | Proportion mediated | Standard covariates | 134.2% | 1.547 | 3.86E-01 |
|  |  |  |  | + 5 sleep traits | 71.1% | 0.431 | 9.92E-02 |
|  | Total sugar (g/day) | 35362 | a (X→M) | Standard covariates | 0.144 | 0.012 | 0.00E+00* |
|  |  |  |  | + 5 sleep traits | 0.139 | 0.012 | 0.00E+00* |
|  |  |  | b (M→Y) | Standard covariates | 6.513 | 0.218 | 0.00E+00* |
|  |  |  |  | + 5 sleep traits | 6.591 | 0.219 | 0.00E+00* |
|  |  |  | c' (X→Y) | Standard covariates | 2.887 | 0.498 | 6.71E-09* |
|  |  |  |  | + 5 sleep traits | 3.073 | 0.500 | 7.87E-10* |
|  |  |  | indirect | Standard covariates | 0.938 | 0.085 | 0.00E+00* |
|  |  |  |  | + 5 sleep traits | 0.919 | 0.086 | 0.00E+00* |
|  |  |  | total | Standard covariates | 3.824 | 0.503 | 2.93E-14* |
|  |  |  |  | + 5 sleep traits | 3.993 | 0.505 | 2.89E-15* |
|  |  |  | Proportion mediated | Standard covariates | 24.5% | 0.036 | 1.61E-11* |
|  |  |  |  | + 5 sleep traits | 23.0% | 0.034 | 6.56E-12* |
| Single item sweet food liking | Free sugar (g/1000kcal) | 35328 | a (X→M) | Standard covariates | 0.256 | 0.024 | 0.00E+00* |
|  |  |  |  | + 5 sleep traits | 0.247 | 0.024 | 0.00E+00* |
|  |  |  | b (M→Y) | Standard covariates | 1.097 | 0.032 | 0.00E+00* |
|  |  |  |  | + 5 sleep traits | 1.087 | 0.032 | 0.00E+00* |
|  |  |  | c' (X→Y) | Standard covariates | 0.333 | 0.145 | 2.12E-02 |
|  |  |  |  | + 5 sleep traits | 0.296 | 0.145 | 4.11E-02 |
|  |  |  | indirect | Standard covariates | 0.281 | 0.028 | 0.00E+00* |
|  |  |  |  | + 5 sleep traits | 0.268 | 0.027 | 0.00E+00* |
|  |  |  | total | Standard covariates | 0.614 | 0.147 | 2.89E-05* |
|  |  |  |  | + 5 sleep traits | 0.565 | 0.147 | 1.26E-04* |
|  |  |  | Proportion mediated | Standard covariates | 45.7% | 0.111 | 3.71E-05* |
|  |  |  |  | + 5 sleep traits | 47.5% | 0.125 | 1.47E-04* |
|  | Free sugar (g/day) | 35331 | a (X→M) | Standard covariates | 0.256 | 0.024 | 0.00E+00* |
|  |  |  |  | + 5 sleep traits | 0.247 | 0.024 | 0.00E+00* |
|  |  |  | b (M→Y) | Standard covariates | 2.865 | 0.078 | 0.00E+00* |
|  |  |  |  | + 5 sleep traits | 2.832 | 0.078 | 0.00E+00* |
|  |  |  | c' (X→Y) | Standard covariates | 2.124 | 0.352 | 1.65E-09* |
|  |  |  |  | + 5 sleep traits | 1.991 | 0.354 | 1.78E-08* |
|  |  |  | indirect | Standard covariates | 0.733 | 0.072 | 0.00E+00* |
|  |  |  |  | + 5 sleep traits | 0.699 | 0.071 | 0.00E+00* |
|  |  |  | total | Standard covariates | 2.857 | 0.358 | 1.55E-15* |
|  |  |  |  | + 5 sleep traits | 2.690 | 0.360 | 7.39E-14* |
|  |  |  | Proportion mediated | Standard covariates | 25.6% | 0.037 | 4.08E-12* |
|  |  |  |  | + 5 sleep traits | 26.0% | 0.040 | 5.35E-11* |
|  | Total sugar (g/1000kcal) | 35328 | a (X→M) | Standard covariates | 0.256 | 0.024 | 0.00E+00* |
|  |  |  |  | + 5 sleep traits | 0.247 | 0.024 | 0.00E+00* |
|  |  |  | b (M→Y) | Standard covariates | 0.724 | 0.043 | 0.00E+00* |
|  |  |  |  | + 5 sleep traits | 0.751 | 0.043 | 0.00E+00* |
|  |  |  | c' (X→Y) | Standard covariates | -0.028 | 0.196 | 8.86E-01 |
|  |  |  |  | + 5 sleep traits | 0.127 | 0.196 | 5.18E-01 |
|  |  |  | indirect | Standard covariates | 0.185 | 0.021 | 0.00E+00* |
|  |  |  |  | + 5 sleep traits | 0.185 | 0.021 | 0.00E+00* |
|  |  |  | total | Standard covariates | 0.157 | 0.197 | 4.24E-01 |
|  |  |  |  | + 5 sleep traits | 0.312 | 0.197 | 1.13E-01 |
|  |  |  | Proportion mediated | Standard covariates | 117.9% | 1.471 | 4.23E-01 |
|  |  |  |  | + 5 sleep traits | 59.4% | 0.375 | 1.14E-01 |
|  | Total sugar (g/day) | 35331 | a (X→M) | Standard covariates | 0.256 | 0.024 | 0.00E+00* |
|  |  |  |  | + 5 sleep traits | 0.247 | 0.024 | 0.00E+00* |
|  |  |  | b (M→Y) | Standard covariates | 2.730 | 0.110 | 0.00E+00* |
|  |  |  |  | + 5 sleep traits | 2.759 | 0.110 | 0.00E+00* |
|  |  |  | c' (X→Y) | Standard covariates | 3.111 | 0.500 | 4.93E-10* |
|  |  |  |  | + 5 sleep traits | 3.297 | 0.502 | 5.13E-11* |
|  |  |  | indirect | Standard covariates | 0.698 | 0.072 | 0.00E+00* |
|  |  |  |  | + 5 sleep traits | 0.681 | 0.072 | 0.00E+00* |
|  |  |  | total | Standard covariates | 3.809 | 0.503 | 3.89E-14* |
|  |  |  |  | + 5 sleep traits | 3.978 | 0.506 | 3.77E-15* |
|  |  |  | Proportion mediated | Standard covariates | 18.3% | 0.029 | 2.01E-10* |
|  |  |  |  | + 5 sleep traits | 17.1% | 0.027 | 1.16E-10* |
| **Sleep predictor (X): Composite sleep score (per unit)** | | | | | | | |
| Average sweet liking (multi-item mean) | Free sugar (g/1000kcal) | 35359 | a (X→M) | Standard covariates | -0.064 | 0.006 | 0.00E+00* |
|  |  |  | b (M→Y) | Standard covariates | 2.625 | 0.063 | 0.00E+00* |
|  |  |  | c' (X→Y) | Standard covariates | -0.402 | 0.067 | 2.29E-09* |
|  |  |  | indirect | Standard covariates | -0.169 | 0.015 | 0.00E+00* |
|  |  |  | total | Standard covariates | -0.571 | 0.069 | 0.00E+00* |
|  |  |  | Proportion mediated | Standard covariates | 29.6% | 0.040 | 1.40E-13* |
|  | Free sugar (g/day) | 35362 | a (X→M) | Standard covariates | -0.064 | 0.006 | 0.00E+00* |
|  |  |  | b (M→Y) | Standard covariates | 6.957 | 0.153 | 0.00E+00* |
|  |  |  | c' (X→Y) | Standard covariates | -1.517 | 0.164 | 0.00E+00* |
|  |  |  | indirect | Standard covariates | -0.447 | 0.041 | 0.00E+00* |
|  |  |  | total | Standard covariates | -1.965 | 0.168 | 0.00E+00* |
|  |  |  | Proportion mediated | Standard covariates | 22.8% | 0.025 | 0.00E+00* |
|  | Total sugar (g/1000kcal) | 35359 | a (X→M) | Standard covariates | -0.064 | 0.006 | 0.00E+00* |
|  |  |  | b (M→Y) | Standard covariates | 1.630 | 0.086 | 0.00E+00* |
|  |  |  | c' (X→Y) | Standard covariates | 0.862 | 0.092 | 0.00E+00* |
|  |  |  | indirect | Standard covariates | -0.105 | 0.011 | 0.00E+00* |
|  |  |  | total | Standard covariates | 0.757 | 0.092 | 2.22E-16* |
|  |  |  | Proportion mediated | Standard covariates | -13.8% | 0.023 | 1.69E-09* |
|  | Total sugar (g/day) | 35362 | a (X→M) | Standard covariates | -0.064 | 0.006 | 0.00E+00* |
|  |  |  | b (M→Y) | Standard covariates | 6.607 | 0.218 | 0.00E+00* |
|  |  |  | c' (X→Y) | Standard covariates | 0.246 | 0.234 | 2.92E-01 |
|  |  |  | indirect | Standard covariates | -0.425 | 0.040 | 0.00E+00* |
|  |  |  | total | Standard covariates | -0.179 | 0.236 | 4.50E-01 |
|  |  |  | Proportion mediated | Standard covariates | 237.9% | 3.123 | 4.46E-01 |
| Single item sweet food liking | Free sugar (g/1000kcal) | 35328 | a (X→M) | Standard covariates | -0.106 | 0.011 | 0.00E+00* |
|  |  |  | b (M→Y) | Standard covariates | 1.090 | 0.032 | 0.00E+00* |
|  |  |  | c' (X→Y) | Standard covariates | -0.457 | 0.068 | 1.54E-11* |
|  |  |  | indirect | Standard covariates | -0.116 | 0.013 | 0.00E+00* |
|  |  |  | total | Standard covariates | -0.573 | 0.069 | 0.00E+00* |
|  |  |  | Proportion mediated | Standard covariates | 20.2% | 0.030 | 1.57E-11* |
|  | Free sugar (g/day) | 35331 | a (X→M) | Standard covariates | -0.106 | 0.011 | 0.00E+00* |
|  |  |  | b (M→Y) | Standard covariates | 2.852 | 0.078 | 0.00E+00* |
|  |  |  | c' (X→Y) | Standard covariates | -1.667 | 0.165 | 0.00E+00* |
|  |  |  | indirect | Standard covariates | -0.303 | 0.033 | 0.00E+00* |
|  |  |  | total | Standard covariates | -1.970 | 0.168 | 0.00E+00* |
|  |  |  | Proportion mediated | Standard covariates | 15.4% | 0.019 | 2.00E-15* |
|  | Total sugar (g/1000kcal) | 35328 | a (X→M) | Standard covariates | -0.106 | 0.011 | 0.00E+00* |
|  |  |  | b (M→Y) | Standard covariates | 0.743 | 0.043 | 0.00E+00* |
|  |  |  | c' (X→Y) | Standard covariates | 0.835 | 0.092 | 0.00E+00* |
|  |  |  | indirect | Standard covariates | -0.079 | 0.010 | 2.22E-16* |
|  |  |  | total | Standard covariates | 0.756 | 0.092 | 2.22E-16* |
|  |  |  | Proportion mediated | Standard covariates | -10.5% | 0.019 | 2.19E-08* |
|  | Total sugar (g/day) | 35331 | a (X→M) | Standard covariates | -0.106 | 0.011 | 0.00E+00* |
|  |  |  | b (M→Y) | Standard covariates | 2.771 | 0.110 | 0.00E+00* |
|  |  |  | c' (X→Y) | Standard covariates | 0.112 | 0.235 | 6.34E-01 |
|  |  |  | indirect | Standard covariates | -0.295 | 0.033 | 0.00E+00* |
|  |  |  | total | Standard covariates | -0.183 | 0.237 | 4.40E-01 |
|  |  |  | Proportion mediated | Standard covariates | 161.2% | 2.072 | 4.36E-01 |

Standard covariates include sex, age, body mass index, socioeconomic status (Townsend Deprivation Index), and ethnicity.
Proportion mediated refers to that mediated by liking, calculated as indirect effect ÷ total effect; estimates >100% or <0% indicate inconsistent mediation, where direct and indirect effects operate in opposite direction.
*p < 0.0025 (Bonferroni-corrected threshold for 20 independent tests)
